# Lower risks of new-onset acute pancreatitis and pancreatic cancer in sodium glucose cotransporter 2 (SGLT2) inhibitors compared to dipeptidyl peptidase-4 (DPP4) inhibitors: a propensity score-matched study with competing risk analysis

**DOI:** 10.1101/2022.05.27.22275702

**Authors:** Oscar Hou In Chou, Jiandong Zhou, Jonathan V Mui, Danish Iltaf Satti, Teddy Tai Loy Lee, Sharen Lee, Edward Christopher Dee, Kenrick Ng, Qingpeng Zhang, Bernard Man Yung Cheung, Fengshi Jing, Gary Tse

## Abstract

**Background:** Dipeptidyl peptidase-4 inhibitors (DPP4I) may be associated with higher risks of acute pancreatitis and pancreatic cancer. This study compared the risks of acute pancreatitis and pancreatic cancer between sodium glucose cotransporter 2 inhibitors (SGLT2I) and DPP4I users.

**Methods:** This was a retrospective population-based cohort study of patients with type-2 diabetes mellitus on either SGLT2I or DPP4I between January 1^st^ 2015 and December 31^st^ 2020 in Hong Kong. The primary outcome was new-onset acute pancreatitis and pancreatic cancer. Propensity score matching (1:1 ratio) using the nearest neighbour search was performed. Univariable and multivariable Cox regressions were applied to identify significant predictors.

**Results:** This cohort included 31609 T2DM patients (median age: 67.4 years old [SD: 12.5]; 53.36% males). 6479 patients (20.49%) used SGLT2I, and 25130 patients (70.50%) used DPP4I. After matching, the incidence of acute pancreatitis was significantly lower in SGLT2I users (incidence rate, IR: 0.6; 95% confidence interval, CI: 0.2-1.4) than in DPP4I (IR: 2.1; CI: 1.3-3.0). The incidence of pancreatic cancer was also lower among SGLT2I users (IR: 1.4; 95% CI: 0.7-2.6 vs. 3.6; 95% CI: 2.6-4.9). SGLT2I was associated with lower risks of acute pancreatitis (hazard ratio, HR: 0.11; 95% CI: 0.02-0.51; P=0.0017) and pancreatic cancer (HR: 0.22; 95% CI: 0.039-0.378; P=0.0003) after adjustments. The results were consistent in the competing risk models and the different matching approaches.

**Conclusions:** SGLT2I may be associated with lower risks of new-onset acute pancreatitis and pancreatic cancer after matching and adjustments, underscoring the need for further evaluation in the prospective setting.

**Key messages:** *What is already known on this topic:* T2DM was associated with higher risks of pancreatic cancer. Meanwhile, second-line anti-diabetic drugs were suggested to reduce the risks of pancreatic cancer, although DPP4I was suggested to be associated with acute pancreatitis.

*What this study adds:* SGLT2I was associated with an 89% lower risk of acute pancreatitis and 78% lower risk of pancreatic cancer than DPP4I users.

*How this study might affect research, practice or policy:* The findings of this study may influence the choice of second-line antidiabetic therapy in T2DM patients in terms of the pancreatic safety profile. This study may inspire more studies on the long-term cancer benefits of SGLT2I.

## Introduction

Pancreatic cancer is currently the seventh leading cause of cancer-related death globally, with the Global Cancer Observatory (GLOBACAN) recording 432,242 associated deaths in 2018 (1). As patients are often asymptomatic until advanced disease, pancreatic cancer remains one of the most lethal cancers despite advances in its detection and awareness (2, 3). The aetiology of pancreatic cancer is still unclear; established risk factors include smoking, obesity, and type 2 diabetes mellitus (T2DM) (2, 4). The relationship between T2DM and pancreatic cancer has been studied by multiple epidemiological studies, with most studies showing an increased risk of pancreatic cancer in relation to diabetes (5, 6). Based on the close relationship between T2DM and pancreatic cancer, increasing attention has turned to the possible association between anti-diabetic medication and pancreatic cancer risk. A recent population-based study in Korea found concordant results, with a reduced risk of pancreatic cancer associated with metformin, thiazolidinedione, and dipeptidyl peptidase-4 inhibitor (DPP4I) and an increased risk of pancreatic cancer associated with insulin and sulfonylureas (7). However, the evidence for newer anti-diabetic agents such as DPP4I and sodium-glucose cotransporter 2 inhibitor (SGLT2I) is comparatively sparse.

Previous studies exploring the relationship between DPP4I use and pancreatic cancer have reported conflicting results. Early studies reported an increased risk of pancreatic cancer with DPP4I use (8, 9), but multiple meta-analyses subsequently found DPP4I use was not associated with an increased risk of pancreatitis or pancreatic cancer (10-12). Meanwhile, there has been limited and inconsistent evidence on the association between SGLT2I use and pancreatic cancer. A recent meta-analysis in 2020 found moderate-quality evidence suggesting no increased risk of acute pancreatitis and very limited evidence suggesting no increased risk of pancreatic cancer associated with SGLT2I (13). However, preclinical studies have suggested that pancreatic carcinomas functionally express SGLT2, and SGLT2I may reduce glucose uptake, thereby reducing tumour cell growth (14, 15). Whilst preclinical studies suggest that SGLT2I may be a promising approach for the preventing or even treating of pancreatic cancer, there is currently insufficient clinical evidence to support this. To our knowledge, little evidence exists that conducts a direct head-to-head comparison between DPP4I and SGLT2I on their associated pancreatic safety in T2DM patients. Therefore, the aim of the present study is to compare the risks of pancreatitis and pancreatic cancer in DPP4I and SGLT2I users using a large cohort of Chinese T2DM patients.

## Methods

### Study design and population

This was a retrospective, territory-wide cohort study of T2DM patients treated with SGLT2I or DPP4I between January 1^st^, 2015, and December 31^st^, 2020 in Hong Kong. Patients were followed up until December 31^st^, 2020, or until death. This study was approved by The Joint Chinese University of Hong Kong–New Territories East Cluster Clinical Research Ethics Committee. The patients were identified from the Clinical Data Analysis and Reporting System (CDARS), a territory-wide database that centralizes patient information from individual local hospitals to establish comprehensive medical data, including clinical characteristics, disease diagnosis, laboratory results, and drug treatment details. The system has been used by both our team and other teams in Hong Kong to conduct comparative studies (16) and recently by our team comparing the cardiovascular outcomes between SGLT2I and DPP4I users (17-19). Patients were excluded based on the following criteria: 1) less than one month of drug exposure; 2) on both DPP4I and SGLT2I, or switched between the two drug classes; 3) died within 30 days at initial drug exposure; 4) less than 18 years old at the start of the study; 5) less than 1 year of drug exposure; 6) pregnancy; 7) without complete demographics 8) without complete HbA1c, fasting glucose, and creatinine tests. Patients with prior pancreatic cancer were excluded. Patients with prior acute pancreatitis were also excluded to ensure the new-onset pancreatitis and pancreatic cancer are due to diabetes instead of recurrent pancreatitis **(Figure 1)**.

**Figure 1.**
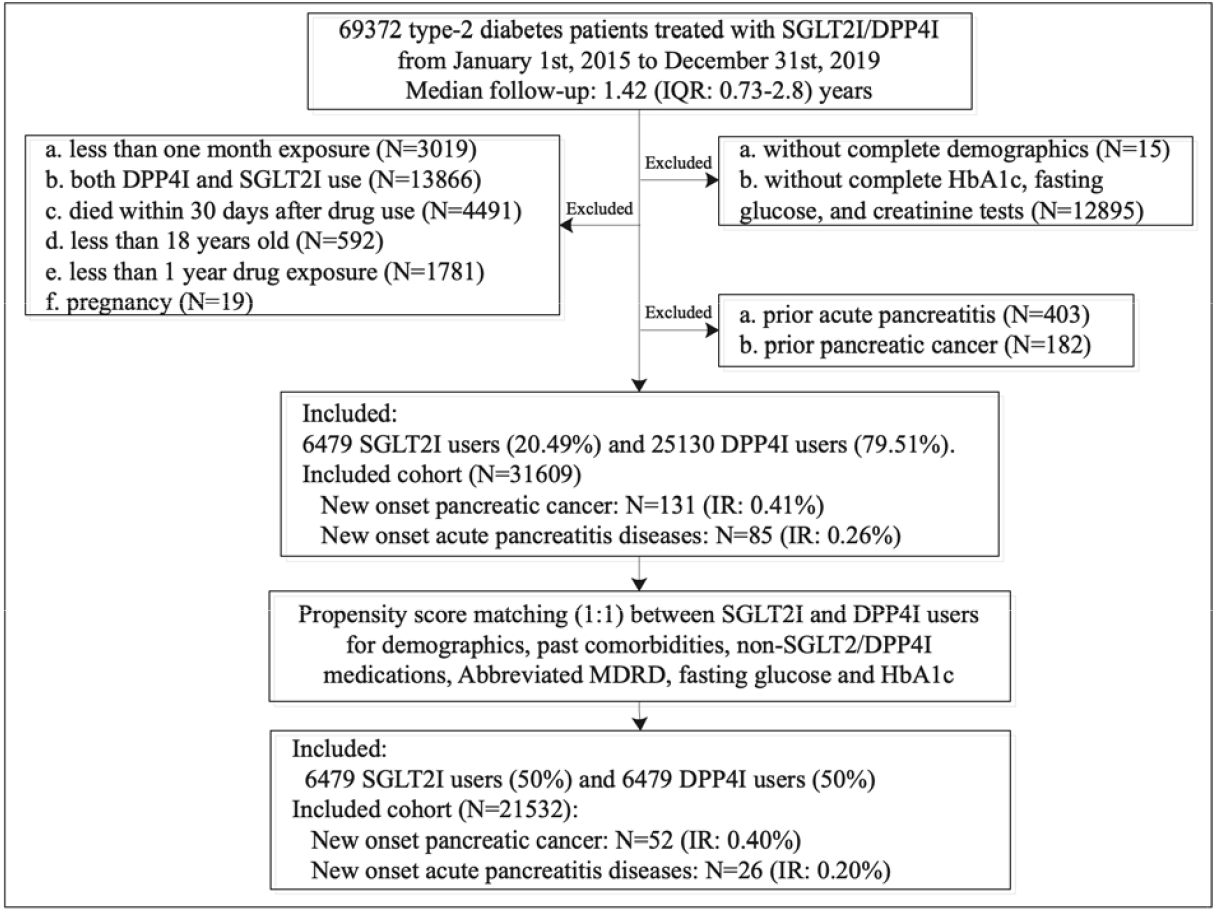
Procedures of data processing for the study cohort. IR: Incidence rate; SGLT2I: Sodium-glucose cotransporter-2 inhibitors; DPP4I: Dipeptidyl peptidase-4 inhibitors.

Patients’ demographics include gender and age of initial drug use (baseline), clinical and biochemical data were extracted for the present study. Prior comorbidities that influenced the treatment selection and the disease outcomes were extracted using the *International Classification of Diseases Ninth Edition* (ICD-9) codes (**Supplementary Table 1**). Charlson’s standard comorbidity index was also calculated. Both cardiovascular medications and anti-diabetic agents were also extracted. The baseline laboratory examinations, including the complete blood count, renal and liver biochemical tests, and the lipid and glucose profiles were extracted. The renal function was calculated using the abbreviated modification of diet in renal disease (MDRD) formula (20).

### Adverse outcomes and statistical analysis

The primary outcomes included new-onset acute pancreatitis (ICD-9: 577.0) and pancreatic cancers (ICD-9: 157.xx). Mortality data were obtained from the Hong Kong Death Registry, a population-based official government registry with the registered death records of all Hong Kong citizens linked to CDARS. The endpoint date of interest for eligible patients was the event presentation date. The endpoint for those without primary outcome presentation was the mortality date or the endpoint of the study (December 31^st^, 2020).

Descriptive statistics are used to summarize baseline clinical and biochemical characteristics of patients with SGLT2I and DPP4I use. For baseline clinical characteristics, the continuous variables were presented as mean (95% confidence interval [CI]/standard deviation [SD])) and the categorical variables were presented as total numbers (percentage). Propensity score matching with 1:1 ratio for SGLT2I use versus DPP4I use based on demographics, Charlson comorbidity index, prior comorbidities, non-SGLT2I/DPP4I medications were performed using the nearest neighbour search strategy. We used Stata software (Version 16.0) to conduct the propensity score matching procedures.

Baseline characteristics between patients with SGLT2I and DPP4I use before and after matching were compared with standardized mean difference (SMD), with SMD<0.20 regarded as well-balanced between two groups. The incidence of acute pancreatitis and pancreatic cancer was derived from dividing the number of outcomes by person-year at risks, which estimate the number of years at risks. Proportional Cox regression models were used to identify significant risk predictors of adverse study outcomes. Cause-specific and subdistribution hazard models were conducted to consider possible competing risks. Multiple propensity adjustment approaches were used, including propensity score stratification (21), propensity score matching with inverse probability of treatment weighting (22) and propensity score matching with stable inverse probability weighting (23). The hazard ratio (HR), 95% CI and P-value were reported. Statistical significance is defined as P-value < 0.05. All statistical analyses were performed with RStudio software (Version: 1.1.456) and Python (Version: 3.6).

## Results

### Basic characteristics

This was a retrospective, territory-wide cohort study of 69372 patients with T2DM treated with SGLT2I/DPP4I between January 1^st^, 2015 and December 31^st^, 2020 in Hong Kong. Patients during the aforementioned period were enrolled and followed up until December 31^st^, 2020 or until their deaths. Patients with less than one month of drug exposure (N=3019), on both DPP4I and SGLT2I (N=13855), died within 30 days at initial drug exposure (N=4491), less than 18 years old at the start of the study (N=592), less than 1 year of drug exposure (N=1781), pregnancy (N=19), without complete demographics or mortality data (N=15), without complete HbA1c, fasting glucose, and creatinine tests (N=12895), prior pancreatitis (N=403) and pancreatic cancer (N=182) were excluded **(Figure 1)**.

After exclusion, this study included a total of 31609 patients with T2DM (median age: 67.4 years old [SD: 12.5]; 53.36% males). 6479 patients (Proportion: 20.49%) used SGLT2Is and 25130 patients (Proportion: 70.50%) used DPP4Is. The DPP4I and SGLT2I cohorts were comparable after matching **(Supplementary Figure 1)**. In the matched cohort, 26 (Proportion: 0.40%) patients developed acute pancreatitis, and 52 patients (Proportion: 0.20%) developed pancreatic cancer. The characteristics of patients are shown in **Table 1, Supplementary Table 3 and 4**.

**Table 1.** Baseline and clinical characteristics of patients with DPP4I v.s. SGLT2I use before and after propensity score matching (1:1). * for SMD≥0.2; SD: standard deviation; CV: coefficient of variation; SCD: sudden cardiac death; VF: ventricular fibrillation; VT: ventricular tachycardia; SGLT2I: sodium glucose cotransporter-2 inhibitor; DPP4I: dipeptidyl peptidase-4 inhibitor; MDRD: modification of diet in renal disease; # indicated the difference between SGLT2I users and DPP4I users.

| Characteristics | Before matching |  |  |  | After matching |  |  |  |
| --- | --- | --- | --- | --- | --- | --- | --- | --- |
|  | All (N=31609)<br>Mean(SD);N or Count(%) | SGLT2I (N=6479)<br>Mean(SD);N or Count(%) | DPP4I (N=25130)<br>Mean(SD);N or Count(%) | SMD<sup>#</sup> | All (N=12958)<br>Mean(SD);N or Count(%) | SGLT2I (N=6479)<br>Mean(SD);N or Count(%) | DPP4I (N=6479)<br>Mean(SD);N or Count(%) | SMD<sup>#</sup> |
| **Adverse outcomes** |  |  |  |  |  |  |  |  |
| New onset pancreatic cancer | 131(0.41%) | 10(0.15%) | 121(0.48%) | 0.06 | 52(0.40%) | 10(0.15%) | 42(0.64%) | 0.08 |
| New onset acute pancreatitis diseases | 85(0.26%) | 4(0.06%) | 81(0.32%) | 0.06 | 26(0.20%) | 4(0.06%) | 22(0.33%) | 0.06 |
| **Demographics** |  |  |  |  |  |  |  |  |
| Male gender | 16869(53.36%) | 3876(59.82%) | 12993(51.70%) | 0.16 | 7605(58.68%) | 3876(59.82%) | 3729(57.55%) | 0.05 |
| Female gender | 14740(46.63%) | 2603(40.17%) | 12137(48.29%) | 0.16 | 5353(41.31%) | 2603(40.17%) | 2750(42.44%) | 0.05 |
| Baseline age, years | 67.4(12.5);n=31609 | 60.6(11.2);n=6479 | 69.2(12.2);n=25130 | 0.73* | 61.9(11.2);n=12958 | 60.6(11.2);n=6479 | 63.2(11.1);n=6479 | 0.19 |
| **Past comorbidities** |  |  |  |  |  |  |  |  |
| Charlson's standard comorbidity index | 2.7(1.7);n=31609 | 1.8(1.2);n=6479 | 2.9(1.8);n=25130 | 0.71* | 1.9(1.3);n=12958 | 1.8(1.2);n=6479 | 2.0(1.3);n=6479 | 0.18 |
| Hypertension | 7640(24.17%) | 1581(24.40%) | 6059(24.11%) | 0.01 | 2999(23.14%) | 1581(24.40%) | 1418(21.88%) | 0.06 |
| Hyperlipidaemia | 1171(3.70%) | 233(3.59%) | 938(3.73%) | 0.01 | 498(3.84%) | 233(3.59%) | 265(4.09%) | 0.03 |
| Heart failure | 533(1.68%) | 87(1.34%) | 446(1.77%) | 0.03 | 175(1.35%) | 87(1.34%) | 88(1.35%) | <0.01 |
| Chronic kidney disease | 11436(36.17%) | 3078(47.50%) | 8358(33.25%) | 0.29* | 5998(46.28%) | 3078(47.50%) | 2920(45.06%) | 0.05 |
| Gallstone | 62(0.19%) | 19(0.29%) | 43(0.17%) | 0.03 | 27(0.20%) | 19(0.29%) | 8(0.12%) | 0.04 |
| Biliary disease | 772(2.44%) | 84(1.29%) | 688(2.73%) | 0.1 | 213(1.64%) | 84(1.29%) | 129(1.99%) | 0.05 |
| Chronic liver disease and cirrhosis | 1032(3.26%) | 277(4.27%) | 755(3.00%) | 0.07 | 534(4.12%) | 277(4.27%) | 257(3.96%) | 0.02 |
| Viral hepatitis | 585(1.85%) | 123(1.89%) | 462(1.83%) | <0.01 | 280(2.16%) | 123(1.89%) | 157(2.42%) | 0.04 |
| History of acute liver injury | 86(0.27%) | 18(0.27%) | 68(0.27%) | <0.01 | 31(0.23%) | 18(0.27%) | 13(0.20%) | 0.02 |
| Other liver disease | 790(2.49%) | 108(1.66%) | 682(2.71%) | 0.07 | 257(1.98%) | 108(1.66%) | 149(2.29%) | 0.05 |
| Stroke/transient ischemic attack | 964(3.04%) | 197(3.04%) | 767(3.05%) | <0.01 | 399(3.07%) | 197(3.04%) | 202(3.11%) | <0.01 |
| Chronic obstructive pulmonary disease | 828(2.61%) | 53(0.81%) | 775(3.08%) | 0.16 | 134(1.03%) | 53(0.81%) | 81(1.25%) | 0.04 |
| Atrial fibrillation | 1444(4.56%) | 162(2.50%) | 1282(5.10%) | 0.14 | 384(2.96%) | 162(2.50%) | 222(3.42%) | 0.05 |
| Ischemic heart disease | 3378(10.68%) | 876(13.52%) | 2502(9.95%) | 0.11 | 1471(11.35%) | 876(13.52%) | 595(9.18%) | 0.14 |
| Peripheral vascular disease | 367(1.16%) | 34(0.52%) | 333(1.32%) | 0.08 | 73(0.56%) | 34(0.52%) | 39(0.60%) | 0.01 |
| Alcoholism or related diagnoses | 119(0.37%) | 12(0.18%) | 107(0.42%) | 0.04 | 29(0.22%) | 12(0.18%) | 17(0.26%) | 0.02 |
| Other cancer except for prior pancreatic cancer | 864(2.73%) | 124(1.91%) | 740(2.94%) | 0.07 | 296(2.28%) | 124(1.91%) | 172(2.65%) | 0.05 |
| **Medications** |  |  |  |  |  |  |  |  |
| SGLT2I duration, days | 518.5(350.3);n=6479 | 518.5(350.3);n=6479 | - | - | 518.5(350.3);n=6479 | 518.5(350.3);n=6479 | - | - |
| DPP4i duration, days | 499.6(278.8);n=25130 | - | 499.6(278.8);n=25130 | - | 522.9(277.3);n=6479 | - | 522.9(277.3);n=6479 | - |
| Metformin | 26037(82.37%) | 5285(81.57%) | 20752(82.57%) | 0.03 | 10916(84.24%) | 5285(81.57%) | 5631(86.91%) | 0.15 |
| Sulphonylurea | 3825(12.10%) | 750(11.57%) | 3075(12.23%) | 0.02 | 1540(11.88%) | 750(11.57%) | 790(12.19%) | 0.02 |
| Acarbose | 316(0.99%) | 75(1.15%) | 241(0.95%) | 0.02 | 155(1.19%) | 75(1.15%) | 80(1.23%) | 0.01 |
| Glucagon-like peptide-1 agonist | 170(0.53%) | 150(2.31%) | 20(0.07%) | 0.21* | 166(1.28%) | 150(2.31%) | 16(0.24%) | 0.18 |
| Other anti-diabetic drugs | 492(1.55%) | 98(1.51%) | 394(1.56%) | <0.01 | 218(1.68%) | 98(1.51%) | 120(1.85%) | 0.03 |
| ACEI/ARB | 2784(8.80%) | 556(8.58%) | 2228(8.86%) | 0.01 | 1192(9.19%) | 556(8.58%) | 636(9.81%) | 0.04 |
| Statins | 31512(99.69%) | 6479(100.00%) | 25033(99.61%) | 0.09 | 12949(99.93%) | 6479(100.00%) | 6470(99.86%) | 0.05 |
| ***Calculated biomarkers*** |  |  |  |  |  |  |  |  |
| Abbreviated MDRD, mL/min/1.73m<sup>2.1</sup> | 76.0(29.1);n=31609 | 90.4(22.4);n=6479 | 72.3(29.5);n=25130 | 0.69* | 89.1(22.8);n=12958 | 90.4(22.4);n=6479 | 87.8(23.1);n=6479 | 0.12 |
| Most severe renal damage (<15 mL/min/1.73m<sup>2</sup>) | 482(1.52%) | 2(0.03%) | 480(1.91%) | 0.19 | 6(0.04%) | 2(0.03%) | 4(0.06%) | 0.01 |
| Severe renal damage ([15, 30] mL/min/1.73m<sup>2</sup>) | 1284(4.06%) | 6(0.09%) | 1278(5.08%) | 0.32* | 15(0.11%) | 6(0.09%) | 9(0.13%) | 0.01 |
| Moderate to severe renal damage ([30, 45] mL/min/1.73m<sup>2</sup>) | 3233(10.22%) | 66(1.01%) | 3167(12.60%) | 0.47* | 189(1.45%) | 66(1.01%) | 123(1.89%) | 0.07 |
| Mild to moderate renal damage ([45, 60] mL/min/1.73m<sup>2</sup>) | 4289(13.56%) | 372(5.74%) | 3917(15.58%) | 0.32* | 899(6.93%) | 372(5.74%) | 527(8.13%) | 0.09 |
| Mild renal damage ([60, 90] mL/min/1.73m<sup>2</sup>) | 12276(38.83%) | 2960(45.68%) | 9316(37.07%) | 0.18 | 5874(45.33%) | 2960(45.68%) | 2914(44.97%) | 0.01 |
| Chronic kidney disease (>90 mL/min/1.73m<sup>2</sup>) | 10045(31.77%) | 3073(47.43%) | 6972(27.74%) | 0.42* | 5975(46.11%) | 3073(47.43%) | 2902(44.79%) | 0.05 |
| Neutrophil-to-lymphocyte ratio | 3.5(4.4);n=15703 | 2.8(2.9);n=3526 | 3.7(4.8);n=12177 | 0.24* | 2.9(3.5);n=6416 | 2.8(2.9);n=3526 | 3.2(4.1);n=2890 | 0.11 |
| Albumin-to-alkaline phosphatase ratio | 0.6(0.2);n=23946 | 0.64(0.2);n=5532 | 0.59(0.2);n=18414 | 0.24* | 0.6(0.2);n=10213 | 0.64(0.2);n=5532 | 0.62(0.2);n=4681 | 0.11 |
| ***Complete blood counts*** |  |  |  |  |  |  |  |  |
| Mean corpuscular volume, fL | 87.4(7.6);n=19283 | 87.0(7.1);n=4409 | 87.5(7.7);n=14874 | 0.06 | 87.0(7.4);n=8010 | 87.04(7.14);n=4409 | 86.98(7.77);n=3601 | 0.01 |
| Basophil, x10<sup>9</sup>/L | 0.0(0.1);n=13543 | 0.04(0.04);n=2666 | 0.04(0.08);n=10877 | 0.02 | 0.0(0.0);n=5262 | 0.04(0.04);n=2666 | 0.04(0.04);n=2596 | 0.01 |
| Eosinophil, x10<sup>9</sup>/L | 0.2(0.3);n=15692 | 0.22(0.2);n=3525 | 0.23(0.28);n=12167 | 0.02 | 0.2(0.3);n=6413 | 0.22(0.2);n=3525 | 0.22(0.37);n=2888 | 0.01 |
| Lymphocyte, x10<sup>9</sup>/L | 2.0(1.0);n=15703 | 2.2(0.8);n=3526 | 1.9(1.0);n=12177 | 0.3* | 2.1(0.9);n=6416 | 2.2(0.8);n=3526 | 2.1(0.9);n=2890 | 0.16 |
| Monocyte, x10<sup>9</sup>/L | 0.5(0.2);n=15703 | 0.53(0.22);n=3526 | 0.53(0.25);n=12177 | 0.02 | 0.5(0.2);n=6416 | 0.53(0.22);n=3526 | 0.51(0.22);n=2890 | 0.09 |
| Neutrophil, x10<sup>9</sup>/L | 5.3(2.8);n=15703 | 5.0(2.3);n=3526 | 5.3(2.9);n=12177 | 0.13 | 5.0(2.4);n=6416 | 5.0(2.3);n=3526 | 5.1(2.5);n=2890 | 0.03 |
| White blood count, x10<sup>9</sup>/L | 8.0(3.1);n=19286 | 7.9(2.5);n=4413 | 8.0(3.2);n=14873 | <0.01 | 7.9(2.7);n=8014 | 7.9(2.5);n=4413 | 7.8(3.0);n=3601 | 0.04 |
| Mean cell haemoglobin, pg | 29.3(3.0);n=19283 | 29.2(2.8);n=4409 | 29.3(3.0);n=14874 | 0.03 | 29.2(3.0);n=8010 | 29.24(2.84);n=4409 | 29.24(3.09);n=3601 | <0.01 |
| Platelet, x10<sup>9</sup>/L | 238.7(72.7);n=19286 | 244.2(67.6);n=4413 | 237.0(74.0);n=14873 | 0.1 | 243.0(70.5);n=8014 | 244.2(67.6);n=4413 | 241.6(73.9);n=3601 | 0.04 |
| Red blood count, x10<sup>12</sup>/L | 4.4(0.7);n=19283 | 4.8(0.6);n=4409 | 4.4(0.7);n=14874 | 0.64* | 4.7(0.6);n=8010 | 4.8(0.6);n=4409 | 4.6(0.6);n=3601 | 0.26* |
| Hematocrit, L/L | 0.4(0.1);n=15864 | 0.41(0.04);n=3712 | 0.38(0.05);n=12152 | 0.7* | 0.4(0.0);n=6643 | 0.41(0.04);n=3712 | 0.4(0.05);n=2931 | 0.32* |
| ***Liver and renal functions*** |  |  |  |  |  |  |  |  |
| K/Potassium, mmol/L | 4.4(0.5);n=31515 | 4.3(0.4);n=6474 | 4.4(0.5);n=25041 | 0.19 | 4.3(0.4);n=12923 | 4.28(0.43);n=6474 | 4.3(0.45);n=6449 | 0.03 |
| Urate, mmol/L | 0.4(0.1);n=4994 | 0.37(0.1);n=1297 | 0.41(0.12);n=3697 | 0.4* | 0.4(0.1);n=2060 | 0.37(0.1);n=1297 | 0.37(0.1);n=763 | 0.05 |
| Albumin, g/L | 41.5(4.1);n=23972 | 42.8(3.2);n=5541 | 41.1(4.2);n=18431 | 0.44* | 42.4(3.5);n=10227 | 42.8(3.2);n=5541 | 42.0(3.7);n=4686 | 0.21* |
| Na/Sodium, mmol/L | 139.3(3.0);n=31532 | 139.4(2.7);n=6475 | 139.3(3.0);n=25057 | 0.04 | 139.4(2.8);n=12926 | 139.4(2.7);n=6475 | 139.3(2.8);n=6451 | 0.04 |
| Urea, mmol/L | 6.9(3.9);n=31530 | 5.6(1.8);n=6469 | 7.2(4.2);n=25061 | 0.5* | 5.6(1.9);n=12928 | 5.6(1.8);n=6469 | 5.7(2.0);n=6459 | 0.05 |
| Protein, g/L | 73.6(5.6);n=22491 | 74.5(4.8);n=5249 | 73.3(5.7);n=17242 | 0.22* | 74.1(5.1);n=9614 | 74.5(4.8);n=5249 | 73.7(5.3);n=4365 | 0.16 |
| Creatinine, umol/L | 101.2(86.5);n=31609 | 77.1(21.0);n=6479 | 107.4(95.4);n=25130 | 0.44* | 78.1(23.6);n=12958 | 77.1(21.0);n=6479 | 79.0(25.9);n=6479 | 0.08 |
| Alkaline phosphatase, U/L | 77.4(32.9);n=24077 | 73.7(25.3);n=5542 | 78.5(34.8);n=18535 | 0.16 | 74.7(27.0);n=10256 | 73.7(25.3);n=5542 | 75.9(28.9);n=4714 | 0.08 |
| Aspartate transaminase, U/L | 27.7(59.8);n=5867 | 27.8(28.6);n=1344 | 27.6(66.3);n=4523 | <0.01 | 28.1(32.2);n=2518 | 27.8(28.6);n=1344 | 28.5(36.0);n=1174 | 0.02 |
| Alanine transaminase, U/L | 27.6(24.7);n=18790 | 34.2(30.7);n=4359 | 25.6(22.2);n=14431 | 0.32* | 31.6(27.9);n=8011 | 34.2(30.7);n=4359 | 28.5(23.7);n=3652 | 0.21* |
| Bilirubin, umol/L | 11.2(6.1);n=23935 | 11.4(6.1);n=5530 | 11.1(6.1);n=18405 | 0.04 | 11.6(6.3);n=10212 | 11.4(6.1);n=5530 | 11.8(6.6);n=4682 | 0.06 |
| ***Lipid and glucose profiles*** |  |  |  |  |  |  |  |  |
| Triglyceride, mmol/L | 1.7(1.4);n=30402 | 1.8(1.7);n=6323 | 1.7(1.3);n=24079 | 0.11 | 1.7(1.5);n=12581 | 1.8(1.7);n=6323 | 1.7(1.4);n=6258 | 0.11 |
| Total cholesterol, mmol/L | 4.0(1.3);n=30422 | 4.2(1.2);n=6326 | 4.0(1.3);n=24096 | 0.17 | 4.1(1.3);n=12586 | 4.2(1.2);n=6326 | 4.0(1.3);n=6260 | 0.11 |
| Low-density lipoprotein, mmol/L | 2.4(0.8);n=27095 | 2.39(0.83);n=5877 | 2.35(0.79);n=21218 | 0.05 | 2.4(0.8);n=11438 | 2.39(0.83);n=5877 | 2.41(0.8);n=5561 | 0.03 |
| High-density lipoprotein, mmol/L | 1.2(0.3);n=27565 | 1.18(0.31);n=5988 | 1.2(0.33);n=21577 | 0.06 | 1.2(0.3);n=11658 | 1.18(0.31);n=5988 | 1.2(0.33);n=5670 | 0.07 |
| Fasting glucose, mmol/L | 8.7(3.7);n=31609 | 9.0(3.8);n=6479 | 8.6(3.7);n=25130 | 0.1 | 8.9(3.6);n=12958 | 9.0(3.8);n=6479 | 8.8(3.5);n=6479 | 0.07 |
| Hemoglobin A1C, % | 8.0(1.6);n=31609 | 8.2(1.6);n=6479 | 7.9(1.5);n=25130 | 0.16 | 8.1(1.5);n=12958 | 8.2(1.6);n=6479 | 8.0(1.5);n=6479 | 0.1 |

### Significant predictors of the study outcomes

Over a follow-up of 19330.3 person-year, 54 cases of pancreatic cancer occurred. Meanwhile, 29 cases of acute pancreatitis occurred during a follow-up of 19341 person-year. Overall, the incidence of pancreatic cancer (IRR: 0.38; 95% CI: 0.19-0.76; P= 0.0048) and acute pancreatitis (IRR: 0.27; 95% CI: 0.09-0.78; P=0.0094) were lower amongst SGLT2I user compared to DPP4I after propensity score matching **(Table 2A)**. Univariable Cox regression identified the significant risk factors for acute pancreatitis and pancreatic cancer before and after propensity score matching (1:1) **(Supplementary Table 5)**. In the multivariable Cox models, SGLT2I was associated with lower risks of acute pancreatitis (HR: 0.11; 95% CI: 0.02-0.51; P=<0.0001) and pancreatic cancer (HR: 0.22; 95% CI: 0.039-0.378; P=0.0003) after adjusting for significant demographics, past comorbidities, non-SGLT2I/DPP4I medications, abbreviated MDRD, fasting glucose, and HbA1c **(Table 2B)**. The cumulative incidence curves stratified by SGLT2I versus DPP4I demonstrated that SGLT2I was associated with a lower cumulative hazard for acute pancreatitis and pancreatic cancer **(Figure 2)**.

**Table 2A.** Annualized incidence rate (IR) per 1000 person-year of new onset pancreatic cancer and new onset acute pancreatitis diseases in the cohort before and after 1:1 propensity score matching.

| Before matching |  |  |  |  | After 1:1 propensity score matching |  |  |  |
| --- | --- | --- | --- | --- | --- | --- | --- | --- |
| New onset pancreatic cancer |  |  |  |  |  |  |  |  |
| Overall | Person-year | Number of events | IR [95% CI] | IRR [95% CI] | Person-year | Number of events | IR [95% CI] | IRR [95% CI] |
|  | 52522.1 | 131 | 2.5[2.1-3] | - | 19330.3 | 54 | 2.8[2.1-3.6] | - |
| DPP4I use | 45359 | 121 | 2.7 [2.2, 3.2] | - | 12167.2 | 44 | 3.6[2.6, 4.9] | - |
| SGLT2I use | 7163.2 | 10 | 1.4[ 0.7, 2.6] | 0.52[0.27, 1.00]* | 7163.2 | 10 | 1.4 [0.7, 2.6] | 0.38[0.19, 0.76]** |
| New onset acute pancreatitis |  |  |  |  |  |  |  |  |
| Overall | Person-year | Number of events | IR [95% CI] | IRR [95% CI] | Person-year | Number of events | IR [95% CI] | IRR [95% CI] |
|  | 52514 | 85 | 1.6[1.3-2] | - | 19341 | 29 | 1.5[1-2.2] | - |
| DPP4I use | 45340.4 | 81 | 1.8 [1.4, 2.2] | - | 12167.5 | 25 | 2.1 [1.3, 3.0] | - |
| SGLT2I use | 7173.5 | 4 | 0.6[0.2, 1.4] | 0.31[0.11, 0.85]* | 7173.5 | 4 | 0.6[0.2, 1.4] | 0.27[0.09,0.78]** |

IR: Incidence rate; IRR: Incidence rate ratio

**Table 2B.** Multivariate Cox regression models with adjustments to predict new onset pancreatic cancer and new onset acute pancreatitis diseases in the matched cohort. * for p≤ 0.05, ** for p ≤ 0.01, *** for p ≤ 0.001; HR: hazard ratio; CI: confidence interval; SGLT2I: sodium glucose cotransporter-2 inhibitor; DPP4I: dipeptidyl peptidase-4 inhibitor. Model 1 adjusted for significant demographics. Model 2 adjusted for significant demographics, and past comorbidities. Model 3 adjusted for significant demographics, past comorbidities, and non-SGLT2I/DPP4I medications. Model 4 adjusted for significant demographics, past comorbidities, non-SGLT2I/DPP4I medications, and abbreviated MDRD. Model 5 adjusted for significant demographics, past comorbidities, non-SGLT2I/DPP4I medications, abbreviated MDRD, fasting glucose, and HbA1c.

|  |  | **Model 1** | **Model 2** | **Model 3** | **Model 4** | **Model 5** |
| --- | --- | --- | --- | --- | --- | --- |
| **SGLT 2I v.s. DPP4I** | **New onset pancreatic cancer HR [95% CI];P value** | 0.41[0.20-0.82];0.0115* | 0.40[0.20-0.81];0.0106* | 0.23[0.040-0.375];0.0002*** | 0.27[0.041-0.392];0.0003*** | 0.22[0.039-0.378];0.0003*** |
|  | **New onset acute pancreatitis HR [95% CI];P value** | 0.31[0.11-0.90];0.0313* | 0.30[0.10-0.88];0.0275* | 0.12[0.03-0.55];<0.0001*** | 0.11[0.02-0.52];<0.0001*** | 0.11[0.02-0.51];<0.0001*** |

**Figure 2.**
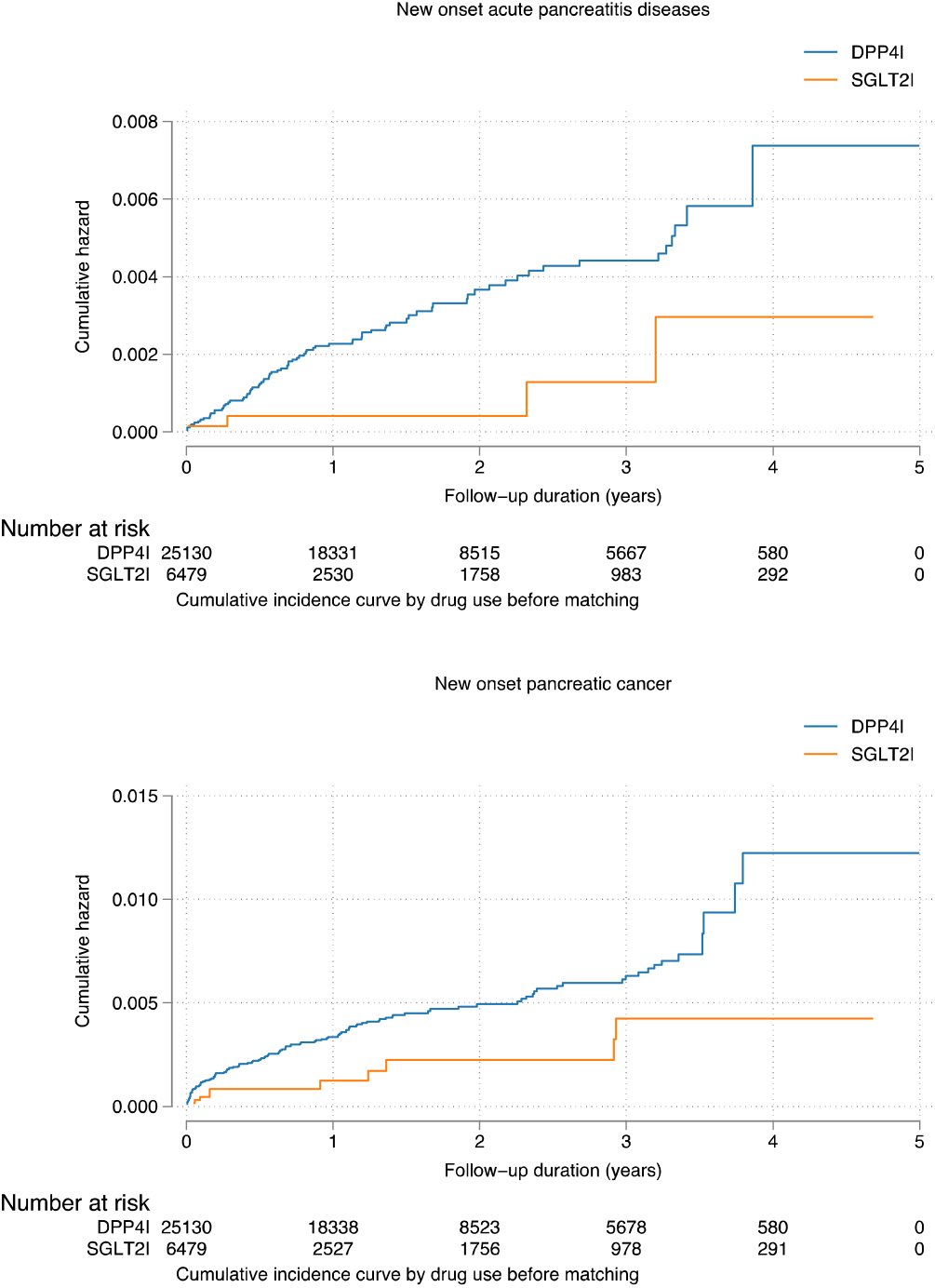
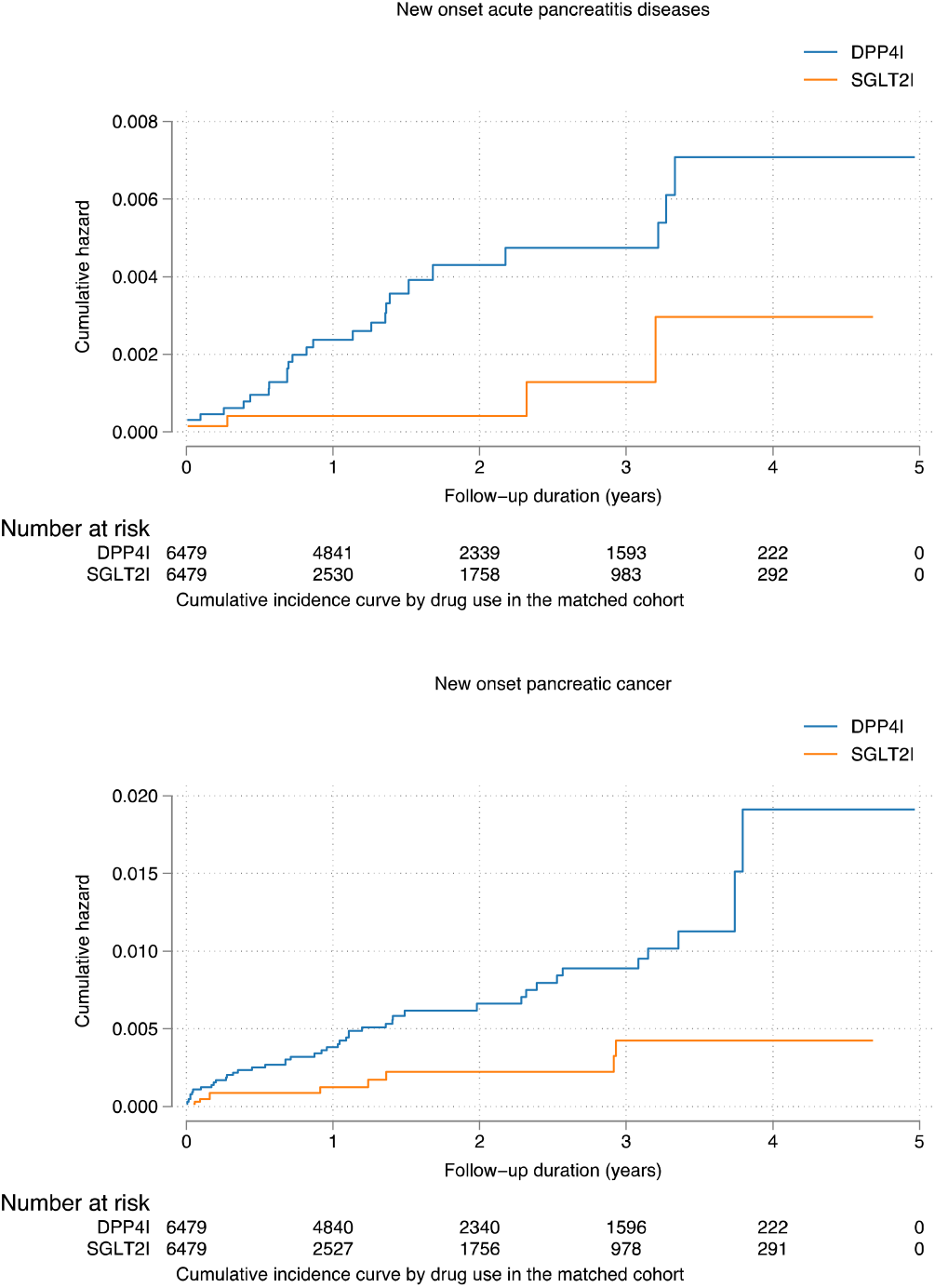
Cumulative incidence curves for new onset pancreatic cancer and new onset acute pancreatitis stratified by drug exposure effects of SGLT2I and DPP4I before and after propensity score matching (1:1).

The sensitivity analyses were performed to confirm the predictiveness of the models. The SGLT2I was associated with lower risks of new-onset acute pancreatitis in the cause-specific hazard (HR: 0.35; 95% CI: 0.12-0.56; P=0.0124) and the subdistribution hazard models (HR: 0.58; 95% CI: 0.15-0.72; P=0.0017). The SGLT2I was also associated with lower risks of new-onset pancreatic cancer in the cause-specific hazard (HR: 0.46; 95% CI: 0.19-0.83; P=0.0023) and the subdistribution hazard models (HR: 0.51; 95% CI: 0.27-0.96; P=0.0047). SGLT2I also was associated with lower risks of new-onset acute pancreatitis and pancreatic cancer across different propensity score approaches **(Supplementary Table 6)**.

## Discussion

In this territory-wide retrospective cohort study, we used real-world data from routine clinical practice to compare the association between SGLT2I versus DPP4I and acute pancreatitis and pancreatic cancer. Our findings demonstrated that SGLT2I was associated with 89% lower risk of acute pancreatitis and 78% lower risk of pancreatic cancer than DPP4I users. To the best of our knowledge, the present study is the first to compare the risks of acute pancreatitis and pancreatic cancer between SGLT2I and DPP4I.

### Comparison with previous studies

Previously, it was suggested that T2DM was associated with pancreatic cancer. This is supported by preclinical studies, which suggest that hyperglycaemia, insulin resistance and pancreatic inflammation are potential mechanisms underlying the relationship between T2DM and pancreatic cancer (24-26). A meta-analysis in 1995 of 20 studies reported a relative risk of pancreatic cancer of 2.1 in diabetic patients compared to non-diabetic patients, while a more recent meta-analysis in 2005 demonstrated an odds ratio for pancreatic cancer of 1.8 (5, 6). As such, anti-diabetic drugs were suggested to reduce the risks of pancreatic cancer. A pooled analysis of 15 case-control studies in 2014 suggested that long-term use of oral anti-diabetic medication is associated with reduced risk of pancreatic cancer while short-term use of insulin is associated with increased risk of pancreatic cancer (27).

Our findings indicated that the use of SGLT2I was associated with lower risks of acute pancreatitis and pancreatic cancer compared to DPP4I. The association of pancreatitis and pancreatic cancer with SGLT2I have been a subject of intense debate, with some studies indicating an increased risk associated with its use, while others were demonstrating a neutral effect (28-32). Most of the documented literature, however, is limited to case reports that lack robust quality evidence. A recent meta-analysis of randomized controlled trials by Tang *et al*. concluded that SGLT2I was not associated with an increased risk of acute pancreatitis or pancreatic cancer (13). Moreover, the meta-regression demonstrated a significantly decreased risk of acute pancreatitis associated with SGLT2I among the patients taking it as monotherapy.

On the other hand, there are conflicting reports regarding the pancreatic safety profile of DPP4I. Previously, the United States Food and Drug Administration Adverse Event Reporting System reported a potential link between DPP4I and acute pancreatitis. However, it was unclear whether DPP4I itself was associated with increased risks of acute pancreatitis, pertaining to the different usage of antidiabetic drugs in the control group (33). Furthermore, multiple meta-analyses have produced contradictory results, with some studies reporting that DPP4I was associated with an increased risk of acute pancreatitis (12, 34, 35), while others suggest no significant increased risks. However, our findings further extend this hypothesis that DPP4I may be associated with pancreatitis and pancreatic cancer, suggesting that its pancreatic safety profile may not be as good as SGLT2I.

While we hypothesized that the protective effects of SGLT2I might be mediated by reducing the number of acute pancreatitis, in our study, we observed that the incidence of acute pancreatitis was lower than pancreatic cancer **(Table 2A)** (36). The same trend was also observed in our previous study regarding the relationship between T2DM and pancreatic cancer **(Data not shown)**. The incidence of acute pancreatitis was similar to that reported in another study in China (37). In Hong Kong, the rate of pancreatic cancer has increased by 90% from 2009 until 2019 (38). This could be partly contributed by the increased incidence of T2DM (39). Meanwhile, in China, the incidence of acute pancreatitis only risen from 30.5 to 39.2 per 100,000 from 2009 to 2014 (40). Furthermore, amongst the excluded patients, only 6 patients with new-onset pancreatic cancer patients had prior pancreatitis. These discrepancies may suggest that while the lower rate of pancreatitis might mediate the lower risks of pancreatic cancer, there might as well be some extra anti-tumour effects associated with SGLT2I as previously suggested.

### Potential underlying mechanisms

Although the precise underlying mechanisms are unclear, several potential explanations exist. It was previously demonstrated in a rat model that treatment with sitagliptin would lead to an increased pancreatic ductal turnover and ductal metaplasia due to the increased level of glucagon-like peptide 1 (GLP-1). GLP-1 was known to stimulate the growth of pancreatic acinar and ductal cells, which would result in ductal occlusion. There are several hypotheses regarding how DPP4I may lead to pancreatic cancer. It was suggested that the pancreatic intraepithelial lesions which precede pancreatic cancer expressed GLP-1. Stimulation of the GLP-1 receptor may trigger increased local replication and proliferation, which gradually leads to pancreatic cancer as the somatic mutation accumulates (41).

SGLT2I has been proposed to have certain anti-tumour benefits, including pancreatic cancer. Scafoglio *et al*. suggested that SGLT2I might play a role in cancer therapy in a xenograft model (14). It has been proposed that SGLT2 is functionally expressed in pancreatic and prostate adenocarcinomas and that SGLT2 inhibitors could block glucose uptake and reduce tumour growth and survival. This might also explain the association of SGLT2I with decreased risk of pancreatic cancer in the present study. It was previously suggested that SGLT2 might promote the progression of pancreatic cancer via the hnRNPK-YAP1 axis, which would enhance YAP1 transcription. Meanwhile, SGLT2I was demonstrated to reverse the action of this pathway (42). Last but not least, SGLT2I was proposed to reduce obesity, which in turns, may reduce the risks of pancreatic cancer (43).

### Clinical implications and the future

Given pancreatic cancer remains one of the rare but most lethal malignancies with a very high mortality to incidence ratio (2), and T2DM is a significant risk factor for this disease (44), there is need to investigate the pancreatic safety profile in SGLT2I and DPP4I. While no conclusions have been reached regarding the relationship between SGLT2I and pancreatic risk, our findings show that SGLT2I may have favourable pancreatic health profiles compared with DPP4I. For instance, glucagon-like peptide-1 receptor agonists was previously suggested to be associated with pancreatic cancer, while later meta-analyses showed no significant increase in risk (45, 46). Furthermore, although no previous cohort study has explored this association specifically with SGLT2I, anti-tumour benefits of several other first-line diabetic medications such as metformin have been well documented in the literature (47). Therefore, by exploring these associations with SGLT2I and DPP4I, we add evidence to the potential anti-tumour role of second-line diabetic medications.

Furthermore, our findings expand on the safety profile of SGLT2I and DPP4I, particularly with regard to acute pancreatitis and pancreatic cancer. In contrast to previous findings, we demonstrated for the first time that SGLT2I might be safe, if not protective, against acute pancreatitis and pancreatic cancer compared with DPP4I. As the T2DM patients may continuously use those newly introduced second-line diabetic medications for a long period of time, our findings may encourage further research in the anti-tumour effects of second-line diabetic medications, owing to the extreme scarcity of the relevant data in the existing literature. The present study used data from routine clinical practice, which may influence the choice of second-line antidiabetic therapy in T2DM patients in terms of the pancreatic risks. Nonetheless, future research exploring the cancer benefits of SGLT2I is warranted.

### Limitations

Several limitations should be noted for the present study. Firstly, given its observational nature, there is inherent information bias due to under-coding, coding errors, and missing data. Secondly, medication adherence can only be assessed indirectly through prescription refills, which are ultimately not a direct measurement of drug exposure. Thirdly, residual, and post-baseline confounding may be present despite robust propensity-matching, particularly with the unavailability of information on cancer risk factors such as smoking, and the potential overlooked alcoholism. Fourthly, the duration of drug exposure has not been controlled for, which may affect their risk against the study outcomes. Furthermore, the follow-up periods were still relatively short despite the statistically significant association was observed. Lastly, our study’s retrospective design necessitates presentation of associations but not causal links between SGLT2I/DPP4I use and the risk of new-onset acute pancreatitis and pancreatic cancer.

## Conclusions

In this real-world cohort study, SGLT2I was associated with lower risks of new-onset acute pancreatitis and pancreatic cancer compared to DPP4I after propensity score matching with adjustments, supporting the need for further evaluation in the prospective setting.

## Supporting information

STROBE

## Data Availability

An anonymised version without identifiable or personal information is available from the corresponding authors upon reasonable request for research purposes.

## Acknowledgements

None.

