## Supplementary Appendix for "Lower risks of new-onset acute pancreatitis and pancreatic cancer in sodium glucose cotransporter 2 (SGLT2) inhibitors compared to dipeptidyl peptidase-4 (DPP4) inhibitors: a propensity score-matched study with competing risk analysis"

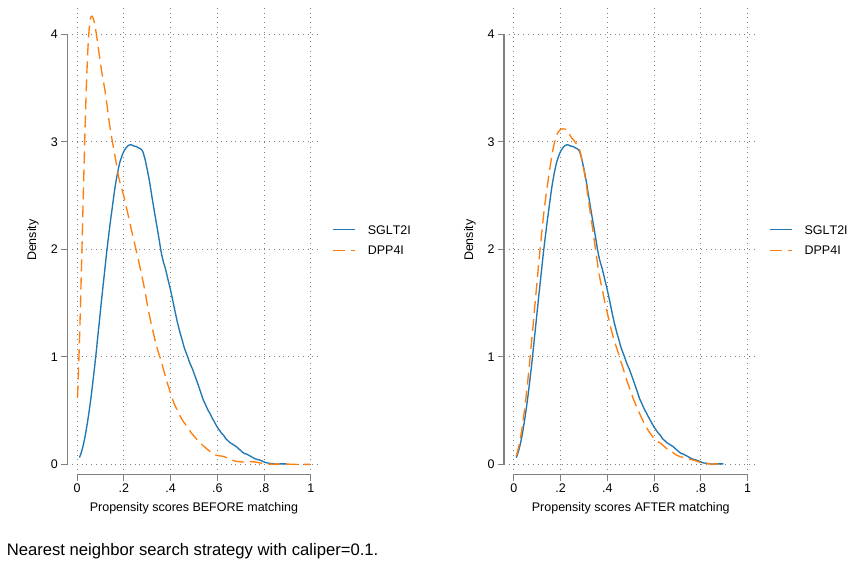

**Supplementary Figure 1. Propensity score matching comparisons and proportional hazard assumption checking with parallel lines for SGLT2I v.s. DPP4I before and after 1:1 matching with nearest neighbor search strategy with caliper of 0.1**

**Supplementary Table 1. ICD9 codes for comorbidities**

Diabetes mellitus 250	250.01	250.02	250.03	250.1	250.11	250.12	250.13	250.2	250.21	250.22	250.23	250.3	250.31	250.32	250.33	250.4	250.41	250.42	250.43	250.5	250.51	250.52	250.53	250.6	250.61	250.62	250.63	250.7	250.71	250.72	250.73	250.8	250.81	250.82	250.83	250.9	250.91	250.92	250.93

Renal diseases	582	582	582.1	582.2	582.4	582.8	582.81	582.89	582.9	583	583	583.1	583.2	583.4	583.6	583.7	585	585.1	585.2	585.3	585.4	585.5	585.6	585.9	586	588	588	588.1	588.8	588.81	588.89	588.9

Acute myocardial infarction 410	410.01	410.02	410.1	410.11	410.12	410.2	410.21	410.22	410.3	410.31	410.32	410.4	410.41	410.42	410.5	410.51	410.52	410.6	410.61	410.62	410.7	410.71	410.72	410.8	410.81	410.82	410.9	410.91	410.92

Heart failure	428	428	428.1	428.2	428.2	428.21	428.22	428.23	428.3	428.3	428.31	428.32	428.33	428.4	428.4	428.41	428.42	428.43	428.9	398.91	402.01	402.11	402.91	404.01	404.03	404.11	404.13	404.91	404.93

Atrial fibrillation	427.31	429.4

Gallstone	574.00	574.01	574.10	574.11	574.20	574.21

Biliary disease	574.30	574.31	574.40	574.41	574.50	574.51	574.60	574.61	574.70	574.71	574.80	574.81	594.90	574.91	575.0	575.10	575.11	575.12	575.2	575.3	575.4	575.5	575.6	575.8	575.9	576.0	576.1	576.2	576.3	576.4	576.5	576.8	576.9

Chronic liver disease and cirrhosis 571.0	571.1	571.2	571.3	571.40	571.41	571.42	471.49	571.5	571.6	571.8,	571.9

Viral hepatitis	070.0	070.1	070.20	070.21	070.22	070.23	070.30	070.31	070.32	070.33	070.41	070.42	070.43	070.44	070.49	070.51	070.52	070.53	070.54	070.59	070.6	070.70	070.71	070.9	573.1	573.2

History of acute liver injury 570	572.2

Other Liver disease	275.1	275.0	572.0	572.4	572.1	572.3	572.8	573.0	573.4	573.8	573.9

Peripheral vascular disease	250.7	443.9	443	443.1	443.2	443.21	443.22	443.23	443.24	443.29	443.8	443.81	443.82	443.89	441	443.9	785.4	V43.4

Stroke/transient ischemic attack	435	435.1	435.2	435.3	435.8	435.9	433.81	433.91	434	436	437	437.1	433.31	433.01	434.01	434.1	434.11	434.9	434.91	437.2	437.3	437.4	437.5	437.6	437.7	437.8	437.9	430	431	432	432.1	432.9

Ischemic heart disease	410.01	410.02	410.1	410.11	410.12	410.2	410.21	410.22	410.3	410.31	410.32	410.4	410.41	410.42	410.5	410.51	410.52	410.6	410.61	410.62	410.7	410.71	410.72	410.8	410.81	410.82	410.9	410.91	410.92	411	411.1	411.8	411.81	411.89	413	413.1	413.9	414	414.01	414.02	414.03	414.04	414.05	414.06	414.07	414.1	414.11	414.12	414.19	414.2	414.3	414.4	414.8	414.9	410	412

Cancer 140	140.1	140.3	140.4	140.5	140.6	140.8	140.9	141	141.1	141.2	141.3	141.4	141.5	141.6	141.8	141.9	142	142.1	142.2	142.8	142.9	143	143.1	143.8	143.9	144	144.1	144.8	144.9	145	145.1	145.2	145.3	145.4	145.5	145.6	145.8	145.9	146	146.1	146.2	146.3	146.4	146.5	146.6	146.7	146.8	146.9	147	147.1	147.2	147.3	147.8	147.9	148	148.1	148.2	148.3	148.8	148.9	149	149.1	149.8	149.9	150	150.1	150.2	150.3	150.4	150.5	150.8	150.9	151	151.1	151.2	151.3	151.4	151.5	151.6	151.8	151.9	152	152.1	152.2	152.3	152.8	152.9	153	153.1	153.2	153.3	153.4	153.5	153.6	153.7	153.8	153.9	154	154.1	154.2	154.3	154.8	155	155.1	155.2	156	156.1	156.2	156.8	156.9	157	157.1	157.2	157.3	157.4	157.8	157.9	158	158.8	158.9	159	159.1	159.8	159.9	160	160.1	160.2	160.3	160.4	160.5	160.8	160.9	161	161.1	161.2	161.3	161.8	161.9	162	162.2	162.3	162.4	162.5	162.8	162.9	163	163.1	163.8	163.9	164	164.1	164.2	164.3	164.8	164.9	165	165.8	165.9	170	170.1	170.2	170.3	170.4	170.5	170.6	170.7	170.8	170.9	171	171.2	171.3	171.4	171.5	171.6	171.7	171.8	171.9	172	172.1	172.2	172.3	172.4	172.5	172.6	172.7	172.8	172.9 173	173.01	173.02	173.09	173.1	173.11	173.12	173.19	173.2	173.21	173.22	173.29	173.3	173.31	173.32	173.39	173.4	173.41	173.42	173.49	173.5	173.51	173.52	173.59	173.6	173.61	173.62	173.69	173.7	173.71	173.72	173.79	173.8	173.81	173.82	173.89	173.9	173.91	173.92	173.99	174	174.1	174.2	174.3	174.4	174.5	174.6	174.8	174.9	175	175.9	176	176.1	176.2	176.3	176.4	176.5	176.8	176.9	179	180	180.1	180.8	180.9	181	182	182.1	182.8	183	183.2	183.3	183.4	183.5	183.8	183.9	184	184.1	184.2	184.3	184.4	184.8	184.9	185	186	186.9	187	187.1	187.2	187.3	187.4	187.5	187.6	187.7	187.8	187.9	188	188.1	188.2	188.3	188.4	188.5	188.6	188.7	188.8	188.9	189	189.1	189.2	189.3	189.4	189.8	189.9	190	190.1	190.2	190.3	190.4	190.5	190.6	190.7	190.8	190.9	191	191.1	191.2	191.3	191.4	191.5	191.6	191.7	191.8	191.9	192	192.1	192.2	192.3	192.8	192.9	193	194	194.1	194.3	194.4	194.5	194.6	194.8	194.9	195	195.1	195.2	195.3	195.4	195.5	195.8	200	200.01	200.02	200.03	200.04	200.05	200.06	200.07	200.08	200.1	200.11	200.12	200.13	200.14	200.15	200.16	200.17	200.18	200.2	200.21	200.22	200.23	200.24	200.25	200.26	200.27	200.28	200.3	200.31	200.32	200.33	200.34	200.35	200.36	200.37	200.38	200.4	200.41	200.42	200.43	200.44	200.45	200.46	200.47	200.48	200.5	200.51	200.52	200.53	200.54	200.55	200.56	200.57	200.58	200.6	200.61	200.62	200.63	200.64	200.65	200.66	200.67	200.68	200.7	200.71	200.72	200.73	200.74	200.75	200.76	200.77	200.78	200.8	200.81	200.82	200.83	200.84	200.85	200.86	200.87	200.88	201	201.01	201.02	201.03	201.04	201.05	201.06	201.07	201.08	201.1	201.11	201.12	201.13	201.14	201.15	201.16	201.17	201.18	201.2	201.21	201.22	201.23	201.24	201.25	201.26	201.27	201.28	201.4	201.41	201.42	201.43	201.44	201.45	201.46	201.47	201.48	201.5	201.51	201.52	201.53	201.54	201.55	201.56	201.57	201.58	201.6	201.61	201.62	201.63	201.64	201.65	201.66	201.67	201.68	201.7	201.71	201.72	201.73	201.74	201.75	201.76	201.77	201.78	201.9	201.91	201.92	201.93	201.94	201.95	201.96	201.97	201.98	202	202.01	202.02	202.03	202.04	202.05	202.06	202.07	202.08	202.1	202.11	202.12	202.13	202.14	202.15	202.16	202.17	202.18	202.2	202.21	202.22	202.23	202.24	202.25	202.26	202.27	202.28	202.3	202.31	202.32	202.33	202.34	202.35	202.36	202.37	202.38	202.4	202.41	202.42	202.43	202.44	202.45	202.46	202.47	202.48	202.5	202.51	202.52	202.53	202.54	202.55	202.56	202.57	202.58	202.6	202.61	202.62	202.63	202.64	202.65	202.66	202.67	202.68	202.7	202.71	202.72	202.73	202.74	202.75	202.76	202.77	202.78	202.8	202.81	202.82	202.83	202.84	202.85	202.86	202.87	202.88	202.9	202.91	202.92	202.93	202.94	202.95	202.96	202.97	202.98	203	203.01	203.02	203.1	203.11	203.12	203.8	203.81	203.82	204	204.01	204.02	204.1	204.11	204.12	204.2	204.21	204.22	204.8	204.81	204.82	204.9	204.91	204.92	205	205.01	205.02	205.1	205.11	205.12	205.2	205.21	205.22	205.3	205.31	205.32	205.8	205.81	205.82	205.9	205.91	205.92	206	206.01	206.02	206.1	206.11	206.12	206.2	206.21	206.22	206.8	206.81	206.82	206.9	206.91	206.92	207	207.01	207.02	207.1	207.11	207.12	207.2	207.21	207.22	207.8	207.81	207.82	208	208.01	208.02	208.1	208.11	208.12	208.2	208.21	208.22	208.8	208.81	208.82	208.9	208.91	208.92 196	196.1	196.2	196.3	196.5	196.6	196.8	196.9	197	197.1	197.2	197.3	197.4	197.5	197.6	197.7	197.8	198	198.1	198.2	198.3	198.4	198.5	198.6	198.7	198.8	198.81	198.82	198.89	199	199.1

Hypertension	401	401.1	401.9	402	402.01	402.1	402.11	402.9	402.91	403	403.01	403.1	403.11	403.9	403.91	404	404.01	404.02	404.03	404.1	404.11	404.12	404.13	404.9	404.91	404.92	404.93	405	405.01	405.09	405.1	405.11	405.19	405.9	405.91	405.99	437.2

Overweight	278	278	278	278.01	278.02	278.03	278.1	278.2	278.3	278.4	278.8

**Supplementary Table 2. Calculations for variability measures**

| **Variability measure** | **Definition** |
| --- | --- |
| Standard deviation | $\sqrt{\frac{1}{Number of measurements}\sum_{i=1}^{Number of measurements} {({test}_{i}-individual mean)}^{2}}$ |
| Coefficient of variation | $\frac{SD}{individual mean}$ |

.

**Supplementary Table 3. Baseline and clinical characteristics of patients with/without new onset pancreatic cancer risk before and after propensity score matching (1:1).**

* for SMD$\geq$0.2; SD: standard deviation; SGLT2I: sodium glucose cotransporter-2 inhibitor; DPP4I: dipeptidyl peptidase-4 inhibitor; CV: coefficient of variation.

|  | **Before matching** |  |  | **After matching** |  |  |
| --- | --- | --- | --- | --- | --- | --- |
| **Characteristics** | **New onset pancreatic cancer (N=131) Mean(SD);N or Count(%)** | **No new onset pancreatic cancer (N=31478) Mean(SD);N or Count(%)** | **SMD** | **New onset pancreatic cancer (N=52) Mean(SD);N or Count(%)** | **No new onset pancreatic cancer (N=12906) Mean(SD);N or Count(%)** | **SMD** |
| ***Demographics*** |  |  |  |  |  |  |
| Male gender | 71(54.19%) | 16798(53.36%) | 0.02 | 34(65.38%) | 7571(58.66%) | 0.14 |
| Female gender | 60(45.80%) | 14680(46.63%) | 0.02 | 18(34.61%) | 5335(41.33%) | 0.14 |
| Baseline age, years | 70.7(10.7);n=131 | 67.4(12.5);n=31478 | 0.29* | 67.1(10.2);n=52 | 61.9(11.2);n=12906 | 0.49* |
| ***Past comorbidities*** |  |  |  |  |  |  |
| Charlson's standard comorbidity index | 3.5(2.3);n=131 | 2.7(1.7);n=31478 | 0.42* | 2.8(2.1);n=52 | 1.9(1.3);n=12906 | 0.49* |
| Hypertension | 35(26.71%) | 7605(24.15%) | 0.06 | 16(30.76%) | 2983(23.11%) | 0.17 |
| Hyperlipidaemia | 3(2.29%) | 1168(3.71%) | 0.08 | 2(3.84%) | 496(3.84%) | <0.01 |
| Heart failure | 1(0.76%) | 532(1.69%) | 0.08 | 1(1.92%) | 174(1.34%) | 0.05 |
| Chronic kidney disease | 38(29.00%) | 11398(36.20%) | 0.15 | 15(28.84%) | 5983(46.35%) | 0.37* |
| Gallstone | 4(3.05%) | 58(0.18%) | 0.23* | 1(1.92%) | 26(0.20%) | 0.17 |
| Bilirary disease | 17(12.97%) | 755(2.39%) | 0.41* | 10(19.23%) | 203(1.57%) | 0.60* |
| Chronic liver disease and cirrhosis | 11(8.39%) | 1021(3.24%) | 0.22* | 5(9.61%) | 529(4.09%) | 0.22* |
| Viral hepatitis | 5(3.81%) | 580(1.84%) | 0.12 | 3(5.76%) | 277(2.14%) | 0.19 |
| History of acute liver injury | 8(6.10%) | 78(0.24%) | 0.34* | 5(9.61%) | 26(0.20%) | 0.45* |
| Other liver disease | 8(6.10%) | 782(2.48%) | 0.18 | 3(5.76%) | 254(1.96%) | 0.2 |
| Stroke/transient ischemic attack | 4(3.05%) | 960(3.04%) | <0.01 | 0(0.00%) | 399(3.09%) | 0.25* |
| Chronic obstructive pulmonary disease | 4(3.05%) | 824(2.61%) | 0.03 | 2(3.84%) | 132(1.02%) | 0.18 |
| Atrial fibrillation | 4(3.05%) | 1440(4.57%) | 0.08 | 1(1.92%) | 383(2.96%) | 0.07 |
| Ischemic heart disease | 12(9.16%) | 3366(10.69%) | 0.05 | 6(11.53%) | 1465(11.35%) | 0.01 |
| Peripheral vascular disease | 1(0.76%) | 366(1.16%) | 0.04 | 0(0.00%) | 73(0.56%) | 0.11 |
| Alcoholism or related diagnoses | 1(0.76%) | 118(0.37%) | 0.05 | 0(0.00%) | 29(0.22%) | 0.07 |
| Other cancer except for prior pancreatic cancer | 5(3.81%) | 859(2.72%) | 0.06 | 1(1.92%) | 295(2.28%) | 0.03 |
| ***Medications*** |  |  |  |  |  |  |
| SGLT2I v.s. DPP4I | 10(7.63%) | 6469(20.55%) | 0.38* | 10(19.23%) | 6469(50.12%) | 0.69* |
| SGLT2I duration, days | 392.4(252.8);n=10 | 518.7(350.4);n=6469 | 0.41* | 392.4(252.8);n=10 | 518.7(350.4);n=6469 | 0.41* |
| DPP4i duration, days | 425.0(327.6);n=121 | 499.9(278.5);n=25009 | 0.25* | 493.5(433.4);n=42 | 523.1(276.0);n=6437 | 0.08 |
| Metformin | 107(81.67%) | 25930(82.37%) | 0.02 | 44(84.61%) | 10872(84.23%) | 0.01 |
| Sulphonylurea | 19(14.50%) | 3806(12.09%) | 0.07 | 9(17.30%) | 1531(11.86%) | 0.15 |
| Acarbose | 3(2.29%) | 313(0.99%) | 0.1 | 2(3.84%) | 153(1.18%) | 0.17 |
| Glucagon-like peptide-1 agonist | 5(3.81%) | 165(0.52%) | 0.23* | 4(7.69%) | 162(1.25%) | 0.32* |
| Other anti-diabetic drugs | 4(3.05%) | 488(1.55%) | 0.1 | 0(0.00%) | 218(1.68%) | 0.19 |
| ACEI/ARB | 11(8.39%) | 2773(8.80%) | 0.01 | 4(7.69%) | 1188(9.20%) | 0.05 |
| Statins | 130(99.23%) | 31382(99.69%) | 0.06 | 52(99.99%) | 12897(99.93%) | 0.04 |
| ***Calculated biomarkers*** |  |  |  |  |  |  |
| Abbreviated MDRD, mL/min/1.73m^2.1 | 72.7(24.6);n=131 | 76.0(29.1);n=31478 | 0.12 | 77.2(22.6);n=52 | 89.2(22.8);n=12906 | 0.53* |
| Most severe renal damage (<15 mL/min/1.73m^2) | 2(1.52%) | 480(1.52%) | <0.01 | 0(0.00%) | 6(0.04%) | 0.03 |
| Severe renal damage ([15, 30) mL/min/1.73m^2) | 4(3.05%) | 1280(4.06%) | 0.05 | 0(0.00%) | 15(0.11%) | 0.05 |
| Moderate to severe renal damage ([30, 45) mL/min/1.73m^2) | 14(10.68%) | 3219(10.22%) | 0.02 | 4(7.69%) | 185(1.43%) | 0.30* |
| Mild to moderate renal damage ([45, 60) mL/min/1.73m^2) | 18(13.74%) | 4271(13.56%) | 0.01 | 9(17.30%) | 890(6.89%) | 0.32* |
| Mild renal damage ([60, 90] mL/min/1.73m^2) | 61(46.56%) | 12215(38.80%) | 0.16 | 24(46.15%) | 5850(45.32%) | 0.02 |
| Chronic kidney disease (>90 mL/min/1.73m^2) | 32(24.42%) | 10013(31.80%) | 0.16 | 15(28.84%) | 5960(46.18%) | 0.36* |
| Neutrophil-to-lymphocyte ratio | 3.9(4.6);n=75 | 3.5(4.4);n=15628 | 0.09 | 3.1(1.8);n=27 | 2.9(3.5);n=6389 | 0.05 |
| Albumin-to-alkaline phosphatase ratio | 0.59(0.23);n=109 | 0.6(0.2);n=23837 | 0.07 | 0.59(0.22);n=46 | 0.63(0.2);n=10167 | 0.18 |
| ***Complete blood counts*** |  |  |  |  |  |  |
| Mean corpuscular volume, fL | 87.5(7.3);n=92 | 87.4(7.6);n=19191 | 0.01 | 88.8(5.5);n=35 | 87.0(7.4);n=7975 | 0.28* |
| Basophil, x10^9/L | 0.04(0.03);n=65 | 0.04(0.07);n=13478 | 0.06 | 0.05(0.03);n=23 | 0.04(0.04);n=5239 | 0.32* |
| Eosinophil, x10^9/L | 0.2(0.16);n=75 | 0.23(0.27);n=15617 | 0.12 | 0.23(0.16);n=27 | 0.22(0.29);n=6386 | 0.05 |
| Lymphocyte, x10^9/L | 1.9(0.8);n=75 | 2.0(1.0);n=15628 | 0.12 | 1.9(0.8);n=27 | 2.1(0.9);n=6389 | 0.25* |
| Monocyte, x10^9/L | 0.54(0.26);n=75 | 0.53(0.25);n=15628 | 0.04 | 0.53(0.22);n=27 | 0.52(0.22);n=6389 | 0.03 |
| Neutrophil, x10^9/L | 5.5(2.6);n=75 | 5.3(2.8);n=15628 | 0.08 | 5.2(2.0);n=27 | 5.0(2.4);n=6389 | 0.06 |
| White blood count, x10^9/L | 7.9(2.5);n=92 | 8.0(3.1);n=19194 | 0.01 | 7.6(1.9);n=35 | 7.9(2.7);n=7979 | 0.11 |
| Mean cell haemoglobin, pg | 29.4(3.0);n=92 | 29.3(3.0);n=19191 | 0.03 | 30.0(2.4);n=35 | 29.2(3.0);n=7975 | 0.27* |
| Platelet, x10^9/L | 235.3(82.2);n=92 | 238.7(72.6);n=19194 | 0.04 | 217.4(81.1);n=35 | 243.1(70.4);n=7979 | 0.34* |
| Red blood count, x10^12/L | 4.42(0.69);n=92 | 4.45(0.7);n=19191 | 0.04 | 4.6(0.6);n=35 | 4.7(0.6);n=7975 | 0.13 |
| Hematocrit, L/L | 0.38(0.05);n=82 | 0.39(0.05);n=15782 | 0.09 | 0.41(0.04);n=27 | 0.4(0.04);n=6616 | 0.03 |
| ***Liver and renal functions*** |  |  |  |  |  |  |
| K/Potassium, mmol/L | 4.3(0.5);n=131 | 4.4(0.5);n=31384 | 0.05 | 4.32(0.45);n=52 | 4.29(0.44);n=12871 | 0.06 |
| Urate, mmol/L | 0.42(0.14);n=17 | 0.4(0.12);n=4977 | 0.15 | 0.42(0.08);n=6 | 0.37(0.1);n=2054 | 0.57* |
| Albumin, g/L | 41.2(3.9);n=109 | 41.5(4.1);n=23863 | 0.08 | 41.4(3.7);n=46 | 42.4(3.5);n=10181 | 0.29* |
| Na/Sodium, mmol/L | 139.1(2.8);n=131 | 139.3(3.0);n=31401 | 0.1 | 139.0(2.8);n=52 | 139.4(2.8);n=12874 | 0.14 |
| Urea, mmol/L | 6.8(3.7);n=131 | 6.9(3.9);n=31399 | 0.02 | 6.3(2.3);n=52 | 5.6(1.9);n=12876 | 0.31* |
| Protein, g/L | 72.7(5.3);n=97 | 73.6(5.6);n=22394 | 0.16 | 72.7(5.6);n=40 | 74.1(5.1);n=9574 | 0.27* |
| Creatinine, umol/L | 105.0(113.7);n=131 | 101.2(86.4);n=31478 | 0.04 | 90.7(30.2);n=52 | 78.0(23.6);n=12906 | 0.47* |
| Alkaline phosphatase, U/L | 84.2(48.7);n=109 | 77.4(32.8);n=23968 | 0.17 | 86.2(61.2);n=46 | 74.7(26.8);n=10210 | 0.24* |
| Aspartate transaminase, U/L | 51.1(88.2);n=39 | 27.5(59.6);n=5828 | 0.31* | 90.8(125.9);n=13 | 27.8(30.8);n=2505 | 0.69* |
| Alanine transaminase, U/L | 34.4(44.4);n=87 | 27.5(24.6);n=18703 | 0.19 | 40.1(38.6);n=30 | 31.6(27.8);n=7981 | 0.25* |
| Bilirubin, umol/L | 14.6(19.2);n=109 | 11.2(6.0);n=23826 | 0.24* | 17.4(28.0);n=46 | 11.5(6.1);n=10166 | 0.29* |
| ***Lipid and glucose profiles*** |  |  |  |  |  |  |
| Triglyceride, mmol/L | 1.7(1.45);n=124 | 1.7(1.39);n=30278 | <0.01 | 1.6(1.1);n=50 | 1.7(1.5);n=12531 | 0.1 |
| Total cholesterol, mmol/L | 3.9(1.6);n=124 | 4.0(1.3);n=30298 | 0.1 | 3.9(1.5);n=50 | 4.1(1.3);n=12536 | 0.18 |
| Low-density lipoprotein, mmol/L | 2.5(0.9);n=105 | 2.4(0.8);n=26990 | 0.14 | 2.5(1.0);n=42 | 2.4(0.8);n=11396 | 0.06 |
| High-density lipoprotein, mmol/L | 1.16(0.39);n=107 | 1.19(0.33);n=27458 | 0.1 | 1.19(0.4);n=43 | 1.19(0.32);n=11615 | 0.01 |
| Fasting glucose, mmol/L | 9.6(4.6);n=131 | 8.7(3.7);n=31478 | 0.21* | 9.9(3.5);n=52 | 8.9(3.6);n=12906 | 0.28* |
| Hemoglobin A1C, % | 8.2(1.7);n=131 | 8.0(1.6);n=31478 | 0.13 | 8.6(1.9);n=52 | 8.1(1.5);n=12906 | 0.28* |

**Supplementary Table 4. Baseline and clinical characteristics of patients with/without new onset acute pancreatitis diseases before and after propensity score matching (1:1).**

* for SMD$\geq$0.2; SD: standard deviation; SGLT2I: sodium glucose cotransporter-2 inhibitor; DPP4I: dipeptidyl peptidase-4 inhibitor; CV: coefficient of variation.

|  | **Before matching** |  |  | **After matching** |  |  |
| --- | --- | --- | --- | --- | --- | --- |
| **Characteristics** | **New onset acute pancreatitis diseases (N=85) Mean(SD);N or Count(%)** | **No new onset acute pancreatitis diseases (N=31524) Mean(SD);N or Count(%)** | **SMD** | **New onset acute pancreatitis diseases (N=26) Mean(SD);N or Count(%)** | **No new onset acute pancreatitis diseases (N=12932) Mean(SD);N or Count(%)** | **SMD** |
| ***Demographics*** |  |  |  |  |  |  |
| Male gender | 45(52.94%) | 16824(53.36%) | 0.01 | 14(53.84%) | 7591(58.69%) | 0.1 |
| Female gender | 40(47.05%) | 14700(46.63%) | 0.01 | 12(46.15%) | 5341(41.30%) | 0.1 |
| Baseline age, years | 70.0(14.7);n=85 | 67.4(12.4);n=31524 | 0.19 | 64.0(10.4);n=26 | 61.9(11.2);n=12932 | 0.2 |
| ***Past comorbidities*** |  |  |  |  |  |  |
| Charlson's standard comorbidity index | 3.0(1.9);n=85 | 2.7(1.7);n=31524 | 0.2* | 2.1(1.4);n=26 | 1.9(1.3);n=12932 | 0.15 |
| Hypertension | 24(28.23%) | 7616(24.15%) | 0.09 | 7(26.92%) | 2992(23.13%) | 0.09 |
| Hyperlipidaemia | 4(4.70%) | 1167(3.70%) | 0.05 | 1(3.84%) | 497(3.84%) | <0.01 |
| Heart failure | 4(4.70%) | 529(1.67%) | 0.17 | 0(0.00%) | 175(1.35%) | 0.17 |
| Chronic kidney disease | 23(27.05%) | 11413(36.20%) | 0.2 | 4(15.38%) | 5994(46.35%) | 0.71* |
| Gallstone | 5(5.88%) | 57(0.18%) | 0.34* | 0(0.00%) | 27(0.20%) | 0.06 |
| Bilirary disease | 8(9.41%) | 764(2.42%) | 0.30* | 1(3.84%) | 212(1.63%) | 0.14 |
| Chronic liver disease and cirrhosis | 7(8.23%) | 1025(3.25%) | 0.22* | 4(15.38%) | 530(4.09%) | 0.39* |
| Viral hepatitis | 5(5.88%) | 580(1.83%) | 0.21* | 1(3.84%) | 279(2.15%) | 0.1 |
| History of acute liver injury | 5(5.88%) | 81(0.25%) | 0.33* | 0(0.00%) | 31(0.23%) | 0.07 |
| Other liver disease | 4(4.70%) | 786(2.49%) | 0.12 | 2(7.69%) | 255(1.97%) | 0.27* |
| Stroke/transient ischemic attack | 2(2.35%) | 962(3.05%) | 0.04 | 1(3.84%) | 398(3.07%) | 0.04 |
| Chronic obstructive pulmonary disease | 6(7.05%) | 822(2.60%) | 0.21* | 1(3.84%) | 133(1.02%) | 0.18 |
| Atrial fibrillation | 4(4.70%) | 1440(4.56%) | 0.01 | 0(0.00%) | 384(2.96%) | 0.25* |
| Ischemic heart disease | 13(15.29%) | 3365(10.67%) | 0.14 | 3(11.53%) | 1468(11.35%) | 0.01 |
| Peripheral vascular disease | 3(3.52%) | 364(1.15%) | 0.16 | 1(3.84%) | 72(0.55%) | 0.23* |
| Alcoholism or related diagnoses | 3(3.52%) | 116(0.36%) | 0.23* | 0(0.00%) | 29(0.22%) | 0.07 |
| Other cancer except for prior pancreatic cancer | 2(2.35%) | 862(2.73%) | 0.02 | 1(3.84%) | 295(2.28%) | 0.09 |
| ***Medications*** |  |  |  |  |  |  |
| SGLT2I v.s. DPP4I | 4(4.70%) | 6475(20.53%) | 0.49* | 4(15.38%) | 6475(50.06%) | 0.80* |
| SGLT2I duration, days | 269.2(336.1);n=4 | 518.7(350.3);n=6475 | 0.73* | 269.2(336.1);n=4 | 518.7(350.3);n=6475 | 0.73* |
| DPP4i duration, days | 448.9(283.8);n=81 | 499.7(278.8);n=25049 | 0.18 | 492.5(324.4);n=22 | 523.0(277.1);n=6457 | 0.1 |
| Metformin | 71(83.52%) | 25966(82.36%) | 0.03 | 22(84.61%) | 10894(84.24%) | 0.01 |
| Sulphonylurea | 13(15.29%) | 3812(12.09%) | 0.09 | 6(23.07%) | 1534(11.86%) | 0.30* |
| Acarbose | 5(5.88%) | 311(0.98%) | 0.27* | 3(11.53%) | 152(1.17%) | 0.43* |
| Glucagon-like peptide-1 agonist | 4(4.70%) | 166(0.52%) | 0.26* | 2(7.69%) | 164(1.26%) | 0.31* |
| Other anti-diabetic drugs | 3(3.52%) | 489(1.55%) | 0.13 | 1(3.84%) | 217(1.67%) | 0.13 |
| ACEI/ARB | 4(4.70%) | 2780(8.81%) | 0.16 | 2(7.69%) | 1190(9.20%) | 0.05 |
| Statins | 84(98.82%) | 31428(99.69%) | 0.1 | 26(99.99%) | 12923(99.93%) | 0.04 |
| ***Calculated biomarkers*** |  |  |  |  |  |  |
| Abbreviated MDRD, mL/min/1.73m^2.1 | 68.7(28.4);n=85 | 76.0(29.1);n=31524 | 0.26* | 76.9(17.5);n=26 | 89.1(22.8);n=12932 | 0.6* |
| Most severe renal damage (<15 mL/min/1.73m^2) | 1(1.17%) | 481(1.52%) | 0.03 | 0(0.00%) | 6(0.04%) | 0.03 |
| Severe renal damage ([15, 30) mL/min/1.73m^2) | 6(7.05%) | 1278(4.05%) | 0.13 | 0(0.00%) | 15(0.11%) | 0.05 |
| Moderate to severe renal damage ([30, 45) mL/min/1.73m^2) | 11(12.94%) | 3222(10.22%) | 0.09 | 0(0.00%) | 189(1.46%) | 0.17 |
| Mild to moderate renal damage ([45, 60) mL/min/1.73m^2) | 13(15.29%) | 4276(13.56%) | 0.05 | 3(11.53%) | 896(6.92%) | 0.16 |
| Mild renal damage ([60, 90] mL/min/1.73m^2) | 35(41.17%) | 12241(38.83%) | 0.05 | 18(69.23%) | 5856(45.28%) | 0.50* |
| Chronic kidney disease (>90 mL/min/1.73m^2) | 19(22.35%) | 10026(31.80%) | 0.21* | 5(19.23%) | 5970(46.16%) | 0.60* |
| Neutrophil-to-lymphocyte ratio | 4.4(6.7);n=54 | 3.5(4.4);n=15649 | 0.15 | 2.7(1.4);n=16 | 2.9(3.5);n=6400 | 0.1 |
| Albumin-to-alkaline phosphatase ratio | 0.59(0.21);n=72 | 0.6(0.2);n=23874 | 0.07 | 0.64(0.25);n=23 | 0.63(0.2);n=10190 | 0.05 |
| ***Complete blood counts*** |  |  |  |  |  |  |
| Mean corpuscular volume, fL | 87.2(9.1);n=61 | 87.4(7.6);n=19222 | 0.02 | 85.5(7.2);n=18 | 87.0(7.4);n=7992 | 0.21* |
| Basophil, x10^9/L | 0.03(0.04);n=44 | 0.04(0.07);n=13499 | 0.11 | 0.02(0.02);n=12 | 0.04(0.04);n=5250 | 0.43* |
| Eosinophil, x10^9/L | 0.24(0.28);n=54 | 0.23(0.27);n=15638 | 0.06 | 0.1(0.2);n=16 | 0.2(0.3);n=6397 | 0.31* |
| Lymphocyte, x10^9/L | 1.98(0.87);n=54 | 1.98(0.95);n=15649 | <0.01 | 2.2(0.9);n=16 | 2.1(0.9);n=6400 | 0.1 |
| Monocyte, x10^9/L | 0.48(0.21);n=54 | 0.53(0.25);n=15649 | 0.22* | 0.4(0.2);n=16 | 0.5(0.2);n=6400 | 0.34* |
| Neutrophil, x10^9/L | 5.6(2.6);n=54 | 5.3(2.8);n=15649 | 0.11 | 5.4(3.2);n=16 | 5.0(2.4);n=6400 | 0.14 |
| White blood count, x10^9/L | 7.9(2.7);n=61 | 8.0(3.1);n=19225 | 0.01 | 8.4(3.6);n=18 | 7.9(2.7);n=7996 | 0.17 |
| Mean cell haemoglobin, pg | 29.28(3.53);n=61 | 29.3(2.98);n=19222 | <0.01 | 28.5(2.8);n=18 | 29.2(3.0);n=7992 | 0.25* |
| Platelet, x10^9/L | 226.3(94.9);n=61 | 238.7(72.6);n=19225 | 0.15 | 227.7(76.7);n=18 | 243.1(70.5);n=7996 | 0.21* |
| Red blood count, x10^12/L | 4.3(0.8);n=61 | 4.4(0.7);n=19222 | 0.25* | 4.66(0.5);n=18 | 4.7(0.61);n=7992 | 0.07 |
| Hematocrit, L/L | 0.37(0.07);n=55 | 0.39(0.05);n=15809 | 0.25* | 0.4(0.05);n=16 | 0.4(0.04);n=6627 | 0.16 |
| ***Liver and renal functions*** |  |  |  |  |  |  |
| K/Potassium, mmol/L | 4.39(0.54);n=85 | 4.35(0.49);n=31430 | 0.07 | 4.31(0.47);n=26 | 4.29(0.44);n=12897 | 0.03 |
| Urate, mmol/L | 0.5(0.1);n=15 | 0.4(0.1);n=4979 | 0.61* | 0.39(0.18);n=2 | 0.37(0.1);n=2058 | 0.13 |
| Albumin, g/L | 41.1(4.4);n=72 | 41.5(4.1);n=23900 | 0.1 | 43.1(3.2);n=23 | 42.4(3.5);n=10204 | 0.21* |
| Na/Sodium, mmol/L | 138.8(3.5);n=85 | 139.3(3.0);n=31447 | 0.17 | 139.2(3.0);n=26 | 139.4(2.8);n=12900 | 0.06 |
| Urea, mmol/L | 7.5(3.7);n=85 | 6.9(3.9);n=31445 | 0.17 | 6.6(1.6);n=26 | 5.6(1.9);n=12902 | 0.55* |
| Protein, g/L | 73.7(6.1);n=65 | 73.6(5.6);n=22426 | 0.02 | 75.6(7.7);n=20 | 74.1(5.1);n=9594 | 0.23* |
| Creatinine, umol/L | 109.8(81.6);n=85 | 101.2(86.5);n=31524 | 0.1 | 85.8(22.4);n=26 | 78.1(23.6);n=12932 | 0.34* |
| Alkaline phosphatase, U/L | 76.9(22.1);n=72 | 77.4(32.9);n=24005 | 0.02 | 73.7(18.9);n=23 | 74.7(27.0);n=10233 | 0.04 |
| Aspartate transaminase, U/L | 22.0(14.0);n=24 | 27.7(59.9);n=5843 | 0.13 | 32.2(20.5);n=8 | 28.1(32.3);n=2510 | 0.15 |
| Alanine transaminase, U/L | 23.9(20.0);n=63 | 27.6(24.7);n=18727 | 0.16 | 27.3(25.9);n=20 | 31.6(27.9);n=7991 | 0.16 |
| Bilirubin, umol/L | 11.3(5.4);n=72 | 11.2(6.1);n=23863 | 0.03 | 12.5(5.3);n=23 | 11.6(6.3);n=10189 | 0.16 |
| ***Lipid and glucose profiles*** |  |  |  |  |  |  |
| Triglyceride, mmol/L | 2.4(4.5);n=84 | 1.7(1.4);n=30318 | 0.21* | 2.9(5.2);n=26 | 1.7(1.5);n=12555 | 0.3* |
| Total cholesterol, mmol/L | 4.1(1.5);n=84 | 4.0(1.3);n=30338 | 0.09 | 4.4(1.4);n=26 | 4.1(1.3);n=12560 | 0.24* |
| Low-density lipoprotein, mmol/L | 2.3(0.8);n=76 | 2.4(0.8);n=27019 | 0.12 | 2.1(0.8);n=25 | 2.4(0.8);n=11413 | 0.35* |
| High-density lipoprotein, mmol/L | 1.1(0.4);n=79 | 1.2(0.3);n=27486 | 0.18 | 1.1(0.4);n=26 | 1.2(0.3);n=11632 | 0.17 |
| Fasting glucose, mmol/L | 8.5(3.2);n=85 | 8.7(3.7);n=31524 | 0.06 | 9.8(3.9);n=26 | 8.9(3.6);n=12932 | 0.23* |
| Hemoglobin A1C, % | 7.98(1.47);n=85 | 7.99(1.56);n=31524 | 0.01 | 8.4(1.5);n=26 | 8.1(1.5);n=12932 | 0.19 |

**Supplementary Table 5. Univariate Cox regression models to predict new onset pancreatic cancer and new onset acute pancreatitis diseases before and after 1:1 propensity score matching.**

* for p≤ 0.05, ** for p ≤ 0.01, *** for p ≤ 0.001; HR: hazard ratio; CI: confidence interval; SD: standard deviation; CV: coefficient of variation; SGLT2I: sodium glucose cotransporter-2 inhibitor; DPP4I: dipeptidyl peptidase-4 inhibitor; MDRD: modification of diet in renal disease.

|  | **Before matching** |  | **After matching** |  |
| --- | --- | --- | --- | --- |
| **Characteristics** | **New onset pancreatic cancer HR [95% CI];P value** | **New onset acute pancreatitis diseases HR [95% CI];P value** | **New onset pancreatic cancer HR [95% CI];P value** | **New onset acute pancreatitis diseases HR [95% CI];P value** |
| ***Demographics*** |  |  |  |  |
| Male gender | 1.07[0.76-1.50];0.7113 | 1.02[0.66-1.56];0.9375 | 1.39[0.79-2.46];0.2567 | 0.87[0.40-1.87];0.7168 |
| Female gender | 1.0[Reference] | 1.0[Reference] | 1.0[Reference] | 1.0[Reference] |
| Baseline age, years | 1.03[1.01-1.04];0.0004*** | 1.02[1.00-1.04];0.0192* | 1.05[1.02-1.08];0.0001*** | 1.02[0.99-1.06];0.1805 |
| ***Past comorbidities*** |  |  |  |  |
| Charlson's standard comorbidity index | 1.30[1.21-1.40];<0.0001*** | 1.17[1.05-1.31];0.0047** | 1.53[1.32-1.77];<0.0001*** | 1.20[0.91-1.58];0.2008 |
| Hypertension | 1.40[0.95-2.07];0.0877 | 1.58[0.98-2.53];0.0606 | 1.87[1.03-3.37];0.0385* | 1.63[0.68-3.89];0.2708 |
| Hyperlipidaemia | 0.64[0.20-2.02];0.4487 | 1.36[0.50-3.71];0.5502 | 1.06[0.26-4.37];0.9338 | 1.05[0.14-7.72];0.9650 |
| Heart failure | 0.56[0.08-3.98];0.5586 | 3.73[1.36-10.19];0.0103* | 1.64[0.23-11.88];0.6237 | - |
| Chronic kidney disease | 0.74[0.51-1.08];0.1230 | 0.68[0.42-1.10];0.1146 | 0.47[0.26-0.86];0.0141* | 0.21[0.07-0.62];0.0044** |
| Gallstone | 17.48[6.46-47.32];<0.0001*** | 37.16[15.05-91.79];<0.0001*** | 11.25[1.55-81.49];0.0166* | - |
| Bilirary disease | 6.33[3.80-10.54];<0.0001*** | 4.44[2.14-9.19];0.0001*** | 14.93[7.49-29.77];<0.0001*** | 2.44[0.33-18.04];0.3807 |
| Chronic liver disease and cirrhosis | 2.87[1.55-5.32];0.0008*** | 2.83[1.31-6.14];0.0083** | 2.46[0.98-6.19];0.0556 | 4.19[1.44-12.15];0.0084** |
| Viral hepatitis | 2.32[0.95-5.68];0.0644 | 3.73[1.51-9.21];0.0043** | 2.95[0.92-9.45];0.0693 | 1.97[0.27-14.54];0.5064 |
| History of acute liver injury | 34.05[16.61-69.81];<0.0001*** | 34.31[13.86-84.92];<0.0001*** | 66.40[26.29-167.72];<0.0001*** | - |
| Other liver disease | 2.77[1.36-5.67];0.0052** | 2.13[0.78-5.80];0.1409 | 3.25[1.01-10.42];0.0477* | 4.42[1.04-18.69];0.0436* |
| Stroke/transient ischemic attack | 1.16[0.43-3.15];0.7642 | 0.92[0.23-3.73];0.9048 | - | 1.53[0.21-11.27];0.6792 |
| Chronic obstructive pulmonary disease | 1.37[0.51-3.72];0.5316 | 3.43[1.49-7.88];0.0037** | 4.02[0.98-16.54];0.0536 | 3.87[0.52-28.59];0.1844 |
| Atrial fibrillation | 0.78[0.29-2.12];0.6276 | 1.26[0.46-3.44];0.6531 | 0.76[0.10-5.50];0.7855 | - |
| Ischemic heart disease | 1.01[0.56-1.83];0.9710 | 1.88[1.04-3.40];0.0366* | 1.27[0.54-2.97];0.5848 | 1.34[0.40-4.46];0.6368 |
| Peripheral vascular disease | 0.79[0.11-5.65];0.8134 | 3.91[1.23-12.39];0.0204* | - | 7.69[1.04-56.81];0.0456* |
| Alcoholism or related diagnoses | 2.23[0.31-15.96];0.4241 | 10.67[3.37-33.79];0.0001*** | - | - |
| Other cancer except for prior pancreatic cancer | 1.74[0.71-4.27];0.2231 | 1.10[0.27-4.48];0.8937 | 0.98[0.14-7.11];0.9855 | 2.03[0.28-15.04];0.4866 |
| ***Medications*** |  |  |  |  |
| SGLT2I v.s. DPP4I | 0.46[0.24-0.88];0.0187* | 0.31[0.11-0.84];0.0214* | 0.36[0.18-0.72];0.0041** | 0.33[0.11-0.97];0.0444* |
| SGLT2I duration, days | 0.998[0.996-1.000];0.0886 | 1.00[0.99-1.00];0.1124 | 0.998[0.996-1.000];0.0886 | 1.00[0.99-1.00];0.1124 |
| DPP4i duration, days | 0.999[0.998-1.000];0.0005*** | 0.999[0.998-1.000];0.0254* | 0.999[0.998-1.000];0.2591 | 0.999[0.998-1.001];0.3987 |
| Metformin | 0.93[0.59-1.44];0.7315 | 1.05[0.59-1.86];0.8734 | 1.03[0.49-2.19];0.9321 | 1.03[0.35-2.99];0.9568 |
| Sulphonylurea | 1.14[0.70-1.85];0.5984 | 1.20[0.67-2.17];0.5400 | 1.35[0.66-2.78];0.4093 | 1.90[0.76-4.74];0.1674 |
| Acarbose | 2.29[0.73-7.20];0.1557 | 6.11[2.48-15.09];0.0001*** | 3.13[0.76-12.86];0.1139 | 10.49[3.15-34.94];0.0001*** |
| Glucagon-like peptide-1 agonist | 5.90[2.39-14.51];0.0001*** | 7.56[2.75-20.77];0.0001*** | 4.71[1.69-13.16];0.0031** | 4.79[1.13-20.34];0.0338* |
| Other anti-diabetic drugs | 1.78[0.66-4.83];0.2557 | 2.10[0.66-6.67];0.2063 | - | 2.27[0.31-16.78];0.4204 |
| ACEI/ARB | 0.95[0.51-1.76];0.8704 | 0.51[0.19-1.39];0.1891 | 0.81[0.29-2.26];0.6938 | 0.81[0.19-3.43];0.7759 |
| Statins | 0.45[0.06-3.22];0.4263 | 0.29[0.04-2.05];0.2130 | - | - |
| ***Calculated biomarkers*** |  |  |  |  |
| Abbreviated MDRD, mL/min/1.73m^2.1 | 1.00[0.99-1.00];0.1960 | 0.99[0.98-1.00];0.0235* | 0.98[0.96-0.99];0.0002*** | 0.98[0.96-0.99];0.0079** |
| Most severe renal damage (<15 mL/min/1.73m^2) | 1.06[0.26-4.27];0.9391 | 0.81[0.11-5.80];0.8311 | - | - |
| Severe renal damage ([15, 30) mL/min/1.73m^2) | 0.75[0.28-2.02];0.5675 | 1.79[0.78-4.10];0.1701 | - | - |
| Moderate to severe renal damage ([30, 45) mL/min/1.73m^2) | 1.03[0.59-1.80];0.9035 | 1.26[0.67-2.38];0.4681 | 5.25[1.89-14.58];0.0014** | - |
| Mild to moderate renal damage ([45, 60) mL/min/1.73m^2) | 1.01[0.61-1.66];0.9700 | 1.14[0.63-2.05];0.6684 | 2.62[1.28-5.37];0.0087** | 1.57[0.47-5.22];0.4635 |
| Mild renal damage ([60, 90] mL/min/1.73m^2) | 1.37[0.97-1.93];0.0745 | 1.10[0.72-1.70];0.6553 | 1.06[0.61-1.82];0.8419 | 2.79[1.21-6.42];0.0156* |
| Chronic kidney disease (>90 mL/min/1.73m^2) | 0.70[0.47-1.04];0.0795 | 0.63[0.38-1.05];0.0737 | 0.47[0.26-0.86];0.0149* | 0.28[0.11-0.74];0.0107* |
| Neutrophil-to-lymphocyte ratio | 1.01[0.98-1.05];0.4022 | 1.02[0.99-1.05];0.1365 | 1.01[0.92-1.11];0.8260 | 0.96[0.77-1.20];0.7283 |
| Albumin-to-alkaline phosphatase ratio | 0.67[0.26-1.74];0.4115 | 0.66[0.21-2.14];0.4930 | 0.36[0.08-1.67];0.1922 | 1.30[0.18-9.17];0.7931 |
| ***Complete blood counts*** |  |  |  |  |
| Mean corpuscular volume, fL | 1.00[0.98-1.03];0.8540 | 1.00[0.97-1.03];0.9135 | 1.04[0.99-1.10];0.1295 | 0.98[0.93-1.03];0.4194 |
| Basophil, x10^9/L | 1.28[0.28-5.83];0.7469 | 0.01[0.00-55.72];0.2931 | 550.90[0.05-5796742.78];0.1816 | 0.00[0.00-1271.73];0.2198 |
| Eosinophil, x10^9/L | 0.56[0.17-1.91];0.3550 | 1.14[0.65-1.99];0.6581 | 1.06[0.45-2.51];0.8883 | 0.04[0.00-2.59];0.1297 |
| Lymphocyte, x10^9/L | 0.85[0.63-1.15];0.2974 | 1.00[0.74-1.36];0.9774 | 0.72[0.43-1.20];0.2074 | 1.10[0.74-1.66];0.6317 |
| Monocyte, x10^9/L | 1.18[0.49-2.82];0.7141 | 0.35[0.09-1.36];0.1293 | 1.07[0.20-5.57];0.9375 | 0.13[0.01-2.26];0.1623 |
| Neutrophil, x10^9/L | 1.03[0.96-1.10];0.4915 | 1.03[0.96-1.12];0.4213 | 1.03[0.88-1.19];0.7497 | 1.07[0.89-1.27];0.4800 |
| White blood count, x10^9/L | 1.00[0.93-1.07];0.9288 | 1.00[0.91-1.09];0.9594 | 0.96[0.83-1.10];0.5529 | 1.04[0.96-1.13];0.3509 |
| Mean cell haemoglobin, pg | 1.01[0.94-1.09];0.7254 | 1.00[0.92-1.09];0.9844 | 1.11[0.97-1.28];0.1221 | 0.94[0.82-1.07];0.3358 |
| Platelet, x10^9/L | 0.999[0.996-1.002];0.6159 | 1.00[0.99-1.00];0.1638 | 0.99[0.99-1.00];0.0247* | 1.00[0.99-1.00];0.3373 |
| Red blood count, x10^12/L | 0.95[0.70-1.27];0.7097 | 0.68[0.47-0.97];0.0358* | 0.83[0.47-1.45];0.5065 | 0.94[0.44-2.02];0.8769 |
| Hematocrit, L/L | 0.18[0.00-10.28];0.4049 | 0.01[0.00-0.74];0.0370* | 2.76[0.00-14085.82];0.8159 | 0.06[0.00-2833.33];0.6156 |
| ***Liver and renal functions*** |  |  |  |  |
| K/Potassium, mmol/L | 0.87[0.61-1.25];0.4569 | 1.10[0.71-1.71];0.6540 | 1.11[0.59-2.06];0.7526 | 1.02[0.42-2.46];0.9694 |
| Urate, mmol/L | 3.57[0.07-170.66];0.5188 | 50.89[1.50-1724.95];0.0288* | 105.10[0.12-89242.09];0.1761 | 17.97[0.00-7502448.95];0.6617 |
| Albumin, g/L | 0.98[0.94-1.02];0.3579 | 0.97[0.92-1.03];0.3268 | 0.93[0.87-1.00];0.0529 | 1.07[0.94-1.21];0.2932 |
| Na/Sodium, mmol/L | 0.97[0.92-1.03];0.2914 | 0.94[0.89-1.01];0.0857 | 0.96[0.87-1.05];0.3567 | 0.99[0.86-1.13];0.8489 |
| Urea, mmol/L | 0.99[0.95-1.04];0.8243 | 1.03[0.99-1.08];0.1243 | 1.11[1.02-1.20];0.0145* | 1.14[1.03-1.25];0.0111* |
| Protein, g/L | 0.97[0.94-1.01];0.1104 | 1.00[0.96-1.05];0.8803 | 0.95[0.89-1.00];0.0697 | 1.06[0.97-1.15];0.1781 |
| Creatinine, umol/L | 1.000[0.999-1.002];0.5764 | 1.001[0.999-1.003];0.3482 | 1.01[1.00-1.01];0.0004*** | 1.01[1.00-1.01];0.1242 |
| Alkaline phosphatase, U/L | 1.00[1.00-1.01];0.0078** | 1.00[0.99-1.01];0.9551 | 1.01[1.00-1.01];0.0035** | 1.00[0.98-1.01];0.8500 |
| Aspartate transaminase, U/L | 1.001[1.000-1.002];0.0865 | 0.99[0.96-1.02];0.4298 | 1.009[1.006-1.013];<0.0001*** | 1.00[0.99-1.02];0.6660 |
| Alanine transaminase, U/L | 1.01[1.00-1.01];0.0028** | 0.99[0.98-1.01];0.2521 | 1.01[1.00-1.01];0.0408* | 0.99[0.97-1.02];0.5178 |
| Bilirubin, umol/L | 1.02[1.02-1.03];<0.0001*** | 1.00[0.97-1.04];0.7916 | 1.02[1.01-1.03];<0.0001*** | 1.01[0.98-1.05];0.4309 |
| ***Lipid and glucose profiles*** |  |  |  |  |
| Triglyceride, mmol/L | 1.00[0.88-1.13];0.9572 | 1.14[1.08-1.21];<0.0001*** | 0.93[0.73-1.18];0.5274 | 1.16[1.07-1.26];0.0004*** |
| Total cholesterol, mmol/L | 0.93[0.81-1.06];0.2656 | 1.08[0.92-1.27];0.3524 | 0.85[0.69-1.06];0.1469 | 1.23[0.91-1.65];0.1725 |
| Low-density lipoprotein, mmol/L | 1.19[0.98-1.45];0.0807 | 0.87[0.64-1.17];0.3548 | 1.12[0.78-1.60];0.5360 | 0.64[0.37-1.11];0.1089 |
| High-density lipoprotein, mmol/L | 0.68[0.37-1.25];0.2154 | 0.52[0.25-1.09];0.0844 | 0.90[0.35-2.30];0.8231 | 0.48[0.13-1.82];0.2783 |
| Fasting glucose, mmol/L | 1.04[1.01-1.07];0.0106* | 0.98[0.92-1.04];0.4677 | 1.05[0.99-1.10];0.0943 | 1.04[0.96-1.12];0.3247 |
| Hemoglobin A1C, % | 1.06[0.96-1.16];0.2663 | 0.96[0.83-1.11];0.6203 | 1.15[0.99-1.34];0.0772 | 1.07[0.85-1.35];0.5774 |

**Supplementary Table 6. Sensitivity analyses for SGLT2I v.s. DPP4I exposure effects predict new onset pancreatic cancer and new onset acute pancreatitis diseases in the matched cohort.**

* for p≤ 0.05, ** for p ≤ 0.01, *** for p ≤ 0.001; SGLT2I: Sodium-glucose cotransporter-2 inhibitors; DPP4I: Dipeptidyl peptidase-4 inhibitors; HR: hazard ratio; CI: confidence interval; PS: propensity score; IPTW: inverse probability of treatment weighting, SIPTW: stable inverse probability of treatment weighting.

| **Model** | **New onset pancreatic cancer**  **HR [95% CI]** | **New onset acute pancreatitis diseases**  **HR [95% CI]** |
| --- | --- | --- |
| Cause-specific hazard models | 0.46[0.19-0.83];0.0023** | 0.35[0.12-0.56];0.0124* |
| Sub-distribution hazard models | 0.51[0.27-0.96];0.0047** | 0.58[0.15-0.72];0.0017** |
| PS stratification | 0.41[0.16-0.64];0.0095** | 0.38[0.12-0.62];0.0108* |
| PS with IPTW | 0.49[0.17-0.76];0.0113* | 0.54[0.15-0.82];0.0087** |
| PS with SIPTW | 0.34[0.25-0.46];0.0142* | 0.42[0.26-0.72];0.0076** |
